# Keeping up with the pathogens: Improved antimicrobial resistance detection and prediction from *Pseudomonas* aeruginosa genomes

**DOI:** 10.1101/2022.08.11.22278689

**Authors:** Danielle E. Madden, Timothy Baird, Scott C. Bell, Kate L. McCarthy, Erin P. Price, Derek S. Sarovich

## Abstract

**Background:** Antimicrobial resistance (AMR) is an intensifying threat that requires urgent mitigation to avoid a post-antibiotic era. The ESKAPE pathogen, *Pseudomonas aeruginosa*, represents one of the greatest AMR concerns due to increasing multi- and pan-drug resistance rates. Shotgun sequencing is quickly gaining traction for *in silico* AMR profiling due to its unambiguity and transferability; however, accurate and comprehensive AMR prediction from *P. aeruginosa* genomes remains an unsolved problem.

**Methods:** We first curated the most comprehensive database yet of known *P. aeruginosa* AMR variants. Next, we performed comparative genomics and microbial genome-wide association study analysis across a Global isolate Dataset (*n*=1877) with paired antimicrobial phenotype and genomic data to identify novel AMR variants. Finally, the performance of our *P. aeruginosa* AMR database, implemented in our ARDaP software, was compared with three previously published *in silico* AMR gene detection or phenotype prediction tools – abritAMR, AMRFinderPlus, ResFinder – across both the Global Dataset and an analysis-naïve Validation Dataset (*n*=102).

**Results:** Our AMR database comprises 3639 mobile AMR genes and 733 AMR-conferring chromosomal variants, including 75 chromosomal variants not previously reported, and 284 chromosomal variants that we show are unlikely to confer AMR. Our pipeline achieved a genotype-phenotype balanced accuracy (bACC) of 85% and 81% across 10 clinically relevant antibiotics when tested against the Global and Validation Datasets, respectively, vs. just 56% and 54% with abritAMR, 58% and 54% with AMRFinderPlus, and 60% and 53% with ResFinder.

**Conclusions:** Our ARDaP software and associated AMR variant database provides the most accurate tool yet for predicting AMR phenotypes in *P. aeruginosa*, far surpassing the performance of current tools. Implementation of our ARDaP-compatible database for routine AMR prediction from *P. aeruginosa* genomes and metagenomes will improve AMR identification, addressing a critical facet in combatting this treatment-refractory pathogen. However, knowledge gaps remain in our understanding of the *P. aeruginosa* resistome, particularly the basis of colistin AMR.

## Background

Antibiotic overuse and misuse [1] has driven the emergence of antimicrobial-resistant (AMR) pathogens globally [2]. We are now on the verge of a ‘post-antibiotic era’, where simple infections threaten to be untreatable with antimicrobials that once revolutionised modern medicine [3]. If unmitigated, AMR infections are predicted to cause 10 million deaths globally by 2050 and cost USD$100 trillion per annum [4].

The ESKAPE pathogen*, Pseudomonas aeruginosa*, represents one of the biggest AMR threats due to its intrinsic resistance towards many antibiotics, environmental ubiquity, ability to infect a wide spectrum of hosts, and high global mortality rate [5–7]. Accurately detecting and predicting AMR phenotype from genotype in *P. aeruginosa* has proven challenging [8], even using machine learning approaches [9], with some approaches as accurate as a coin flip [8]. A major shortcoming of current *in silico* AMR tools is that they largely focus on detecting AMR gene gain [10, 11] and a small number of chromosomally encoded single-nucleotide polymorphisms (SNPs) [12–14]. However, *P. aeruginosa* can also evolve AMR through chromosomal insertions-deletions (indels), loss-of-function mutations (e.g. large deletions or frameshift mutations), structural variants, and copy-number variations (CNVs) [15]. Despite recent advances [11, 12], most AMR tools remain limited in their scope and accuracy [16] – for example, loss-of-function mutations, a major contributor to AMR, are largely ignored [14], AMR databases are often not species-specific [14, 17], they do not resolve to the individual antibiotic level [11], and precursor mutations conferring reduced antimicrobial susceptibility are overlooked. These limitations are especially problematic for accurate AMR detection and prediction in pathogens encoding complex resistomes like *P. aeruginosa* [8].

To address this gap, we curated and validated the most comprehensive *P. aeruginosa*-specific AMR variant database yet, which, when used in conjunction with the Antimicrobial Resistance Detection and Prediction (ARDaP) software [14], enables high-accuracy AMR prediction from *P. aeruginosa* genomes. Performance of our ARDaP-compatible AMR variant database was first assessed across 1877 diverse *P. aeruginosa* strains (“Global Dataset”), and subsequently, across 102 analysis-naïve *P. aeruginosa* strains (“Validation Dataset”). Our approach, which we demonstrate far exceeds the performance to current AMR prediction software, provides a crucial steppingstone towards the routine clinical use of genomics and metagenomics to inform personalised *P. aeruginosa* antimicrobial treatment regimens.

## Methods

### *P. aeruginosa* AMR variant database construction

To cover the spectrum of AMR variants found in *P. aeruginosa*, an ARDaP v1.9 [14] compatible SQLite database was populated with all known AMR variants described in this pathogen to date (https://github.com/dsarov/ARDaP/blob/master/Databases/Pseudomonas_aeruginosa_pao1/). Gene names, locus tags, and genomic coordinates in our AMR variant database were based on PAO1 (NC_002516.2) [18]. An exhaustive literature search was conducted to encompass biomedical literature published from 1980 to April 20^th^ 2023 within the MEDLINE database. Search terms included “antimicrobial resistance” and *“P. aeruginosa”* accompanied by a variable search term for targeted literature, and included either: i) antibiotic class (e.g. carbapenem, aminoglycoside); ii) AMR mechanism (e.g. efflux, gene expression enzymatic inactivation); or iii) a known AMR gene (e.g. *oprD*, *ampC*, *gyrA*).

The resultant *P. aeruginosa* AMR database (Dataset 1) consists of three tables. The first table, ‘1. Antibiotics’, lists the ten clinically relevant antibiotics that were interrogated in our study: amikacin, cefepime, ceftazidime, ciprofloxacin, colistin, imipenem, meropenem, piperacillin, piperacillin/tazobactam, and tobramycin, whether these drugs are first-line, second-line or tertiary, and their associated antibiotic class. The second (‘2. Variants_SNP_indel’) and third (‘3. Coverage’) tables list the entire *P. aeruginosa* mutational resistome [19, 20], which includes all known genetic alterations that can lead to AMR, including the functional loss of chromosomal genes (under, ‘3. Coverage’). These databases include all AMR variants and genes that: i) cause efflux pump upregulation [21]; ii) alter outer membrane permeability [22]; iii) de-repress or alter the substrate range of the AmpC cephalosporinase [23]; and/or iv) alter the antimicrobial target [24]. AMR gene acquisition was interrogated with ResFinder v4.0 [25], using default parameters and retaining only genes identified at 100% similarity and with 100% coverage. Resfinder is integrated within the ARDaP tool and run as part of the default pipeline. The default ResFinder database was manually curated to: 1) remove false-positive hits that were consistently identified in AMR sensitive strains; *bla_OXA-_*_395_1_AY306133_, *bla*_OXA-396_1_AY306134_, *bla*_PAO_4_AY083592_, *bla*_PAO_1_AY083595_, *bla*_PAO_3_FJ666073_, *bla*_PAO_2_FJ666065_, and *crpP*_HM560971; 2) include additional gene variants identified within the Global Dataset in aminoglycoside AMR-conferring genes *aac*(6’)-Ib, *aac*(6’)-IIa, *aac*(6’)-Ib-cr, and *aac*(3)-IIIa; and 3) as per ResFinder recommendations for *P. aeruginosa*, the substrate range for *bla_OXA-2_1_DQ112222_* and *bla_OXA-_*_2_2_GQ466184_ was expanded to include meropenem and imipenem.

For the aminoglycosides (i.e. amikacin and tobramycin), known AMR variants consisted of point mutations in *algA* [26]*, amgS* [27], *fusA1* [26, 28–33], *rplB* [34]*, ptsP* [33, 35], and *tuf1* [26], and loss-of-function mutations in the *nuo* [36–38] pathway. For the carbapenems (i.e. imipenem and meropenem), cephalosporins (i.e. ceftazidime), and β-lactams with or without an inhibitor (i.e. piperacillin and piperacillin/tazobactam), mutations that alter or expand the substrate range of the *ampC* cephalosporinase [23, 39–42] or that alter *ampC* expression, including inactivation of penicillin-binding-protein 4 (PBP4; *dacB*) [43–45], *ampD* [45–47], *ampDh2* [47], *ampDh3* [47], *ampE* [47], *ampR* [48, 49], *mpl* [50], or *ampC-ampR* intergenic region [51], mutations in PBP3 (*ftsI*) [48, 52, 53], and mutations that cause *oprD* loss, inactivation, or down-regulation [49, 54–60], were included. For ciprofloxacin AMR prediction, we included point mutations in *gyrA* [61–67], *gyrB* [62, 63, 65, 68], *parC* [61–63, 66], and *parE* [62, 63, 65]. For colistin AMR prediction, point mutations in *cprS* [52], *pmrA* [69], *pmrB* [31, 69–74], *phoP* [69], and *phoQ* [69] were included.

Efflux pumps play a key role in AMR development in *P. aeruginosa*, predominantly through regulator alterations, which drive efflux pump overexpression [15]. To predict MexAB-OprM upregulation, which is associated with β-lactam (including carbapenem) AMR [21], we included mutations in the *cis* (*mexR* [29, 61, 64, 75, 76]) and *trans* (*nalC* [77, 78] and *nalD* [23, 71, 77–79]) regulators of this efflux pump. For MexCD-OprJ upregulation, which is associated with fluoroquinolone and specific β-lactam AMR [21], mutations and loss-of coverage in the single known regulator, *nfxB* [61, 64] were included. For MexEF-OprN upregulation, which is linked to fluoroquinolone AMR [21], we included function-altering mutations or loss of coverage in *mexS* [80], the LysR family regulator *mexT* [81], and the global regulator *mvaT* [82]. For MexXY upregulation, which is associated with aminoglycoside and fluoroquinolone AMR, we included loss-of-coverage in two refined regions of *mexZ* [83, 84] and the intergenic region (*mexOZ*) between *mexZ* and *mexX* [85, 86]. Additionally, functional gene loss variants across the entire *P. aeruginosa* chromosome, identified via transposon insertion experiments [36, 38, 87], were included.

In addition to variants that confer AMR, mutations in essential AMR-conferring genes that cause unusual antimicrobial susceptibility were included: efflux pumps *mexAB* [88, 89], *mexXY* [90], *mexCD* [91] and *mexEF* [82, 92], and the *ampC* regulators *ampP* and *ampG*, which are required for high-level *ampC* expression [51]. We also included additional targets for (meta)genome quality control and *P. aeruginosa* speciation (*ecfX* [93]), and genes responsible for conferring a hypermutator phenotype (*mutS*, *mutL* and *uvrD* [94]).

### Global Dataset

To comprehensively capture geographic, genomic, and phenotypic diversity, we collated a large and comprehensive *P. aeruginosa* isolate collection (*n=*1877) of all publicly available strains with paired antimicrobial phenotype and genomic data [9, 20, 32, 54, 95–103] (Dataset 2, Table S1). This dataset includes many antimicrobial-susceptible strains, which are essential for developing and refining high-quality AMR databases [14, 104]. Isolates with minimum inhibitory concentration (MIC) data were reclassified as sensitive, intermediate, or resistant using the CLSI M100S-Ed32:2022 guidelines.

### Validation Dataset

To independently evaluate AMR software performance, we examined 102 phylogenetically diverse, analysis-naïve, clinical *P. aeruginosa* strains (Figures S1 and S2). Isolates were obtained from people admitted to hospitals in Qld, Australia, with cystic fibrosis (CF; *n*=42), bacteraemia (*n*=35), chronic obstructive pulmonary disease (COPD; *n*=21), bronchiectasis (*n*=1), ear infection (*n*=1), ulcer (*n*=1), or urinary tract infection (*n*=1), between 2008 and 2020 (Table S2). Eighty-four isolates have previously been genome-sequenced (NCBI BioProject PRJNA761496; GenBank accessions NSXK00000000.1 and NSZK00000000.1) and some have previously undergone antimicrobial susceptibility testing [105, 106]. For the current study, 18 additional COPD isolates (SCHI0038.S.1, SCHI0050.S.3, SCHI0058.S.1, SCHI0058.S.2, SCHI0059.S.1, SCHI0064.S.1, SCHI0065.S.1, SCHI0068.S.3, SCHI0070.S.1, SCHI0070.S.1, SCHI0084.S.1, SCHI0098.S.1, SCHI0103.S.1, SCHI0107.S.1, SCHI0109.S.1, SCHI0109.S.2, SCHI0112.S.1, SCHI0112.S.2) were sequenced and appended to BioProject PRJNA761496. Antimicrobial susceptibility profiles for 8 of 10 clinically-relevant antibiotics were determined across the Validation Dataset (Table S2) using disc diffusions, following CLSI M100S-Ed32:2022 guidelines. Meropenem and ciprofloxacin MICs were determined by ETEST (bioMérieux, Murarrie, Australia) using sensitive, intermediate, and resistant MIC cut-offs of ≤4, 8, and ≥16 µg/mL for meropenem, and ≤0.5, 1, and ≥2 µg/mL for ciprofloxacin. PAO1 (Belgian Coordinated Collection of Microorganisms [BCCM], Ghent University, Belgium) and LMG 6395 (BCCM) were included as antimicrobial-susceptible controls.

### Microbial genome-wide association study (mGWAS) and machine learning (ML) for AMR prediction

To identify novel AMR variants, mGWAS [107] was performed on the Global *P. aeruginosa* Dataset (*n*=1877 strains), with SNPs and indels identified using SPANDx v4.0.1 [108]. To increase the signal-to-noise ratio, variants found in antimicrobial-sensitive isolates were penalised four-fold compared with AMR strains due to a presumed large effect size [109]. The top 50 variants associated with each AMR phenotype were assessed for their ability to improve phenotype prediction; those that improved phenotype prediction were included in the AMR database. Additionally, a supervised ML approach was performed using the Global Dataset and the AMR database as features for model creation.

### Comparative genomic analysis

To identify additional novel AMR variants, we conducted a comparative genomic analysis using SPANDx, with a focus on AMR strains that did not encode a known AMR variant (i.e. false negatives). These strains were compared to their closest antimicrobial-sensitive relative(s) as determined by the whole-genome phylogenetic analysis (**Figure S1**). SNPs and indels that separated AMR from antimicrobial-sensitive strain/s were identified, annotated and investigated with manual inspection to prioritise mutations in known AMR genes. Candidate variants were then tested against the Global Dataset to determine whether they improved phenotype prediction. AMR variants that increased balanced accuracy (bACC) were included in the database; those that did not alter, or that decreased bACC, were discarded.

### AMR prediction analysis

AMR prediction was performed using our *P. aeruginosa* AMR database (v1.0), implemented in ARDaP [110]. ARDaP was chosen as it is the only AMR software that can detect all mutation types (i.e. SNPs, indels, gene gain, gene loss, frameshift mutations, structural variants, and CNVs) [14]. ARDaP also has a built-in feature that automatically generates a clinician-friendly antimicrobial susceptibility summary report for each strain (Figure S3) to simplify *in silico* AMR interpretation [14]. ARDaP performance was compared against four tools for AMR phenotype prediction and/or AMR variant identification: abritAMR [111], RGI v5.1.0 and CARD v3.0.9 [11], ResFinder v4.1 [10], and AMRFinderPlus v3.8.28 [112]. As abritAMR and AMRFinderPlus frequently report predicted AMR phenotypes to the antibiotic class level only, we chose to interpret AMR variant presence for a given class as conferring AMR towards all antibiotics within that class. Importantly, AMRFinderPlus is not intended for clinical phenotype prediction (https://github.com/ncbi/amr/wiki/Interpreting-results#genotype-vs-phenotype) but has been included as a benchmark for gene detection accuracy. For the purposes of software comparisons, gene identification by AMRFinderPlus was interpreted as conferring phenotypic resistance.

A variant scoring scheme has previously been described by Cortes-Lara and colleagues, which employed a 0 (no effect) through 1 (EUCAST AMR) scale to predict *in silico* AMR profiles [102]. We expanded upon this scheme by providing an automated weighted score for all AMR variants in our database that quantifies their contribution, positive or negative, towards AMR development (Dataset 1, ‘Threshold’ column); this score is recorded for each antibiotic on ARDaP’s automatically generated clinician-friendly report (Figure S3), unlike the Cortes-Lara scheme, which requires manual scoring for each antibiotic and strain [102]. Using our scoring system, variants known to cause AMR in isolation score as 100%, whereas AMR variants known to confer AMR in a stepwise manner (that is, only when in combination with other variant/s), or that only result in intermediate resistance, are given a lower score (e.g. 40-50%). This method accounts for both the additive nature of chromosomal mutations in *P. aeruginosa*, and for the decreased AMR potential caused by loss of efflux pumps or essential transcriptional regulators. Acquired AMR genes identified by Resfinder within ARDaP were considered to confer full AMR, i.e. are scored at 100% for the purposes of phenotype prediction.

### Intermediate resistance prediction

The capacity of our AMR variant database to predict intermediate resistance phenotypes was examined against the Global Dataset using the following criteria:

- A true-positive prediction occurred when either: i) an AMR strain was classed as AMR, or ii) an intermediate strain was identified as intermediate;
- A true-negative prediction occurred when an antimicrobial-sensitive strain was identified as antimicrobial sensitive;
- A false-positive prediction occurred when either: i) an antimicrobial-sensitive strain was classed as intermediate or AMR, or ii) an intermediate strain was classed as AMR; and
- A false-negative prediction occurred when either: i) an intermediate strain was classed as antimicrobial-sensitive, or when an AMR strain was predicted to be antimicrobial-sensitive or intermediate.

These rules provided the strictest evaluation criteria for the assessment of ARDaP’s ability to identify intermediate strains. Only ARDaP’s intermediate resistance prediction performance was assessed as abritAMR, AMRFinderPlus, CARD, and ResFinder all lack the capacity to identify intermediate resistance.

### AMR software predictive performance in *P. aeruginosa*

Due to its inability to predict AMR towards individual antibiotics, and a very high rate of false-positive predictions in the Global Dataset, CARD was deemed unsuitable for *P. aeruginosa* AMR analysis and was excluded from further assessment. For all other tools, predictive performance was determined using bACC [113, 114], which averages sensitivity [i.e. true positives/(true positives + false negatives)], and specificity [i.e. true negatives/(true negatives + false positives)]. This metric was chosen as it accounts for dataset imbalance; that is, it minimises over- or under-representation of antimicrobial-sensitive or AMR strains that may otherwise make certain tools appear better or worse due to inherent dataset bias [8]. Additionally, we compared recall (AMR) [true positives/(true positives + false negatives], precision (AMR) or positive predictive value (PPV) [true positives/(true positives + false positives), recall (sensitivity) [true negatives/(true negatives + false positives) and precision (sensitivity) or negative predictive value (NPV) [true negatives/(true negatives + false negatives)] across all software tools.

## Results

### *P. aeruginosa* AMR variant identification and refinement

An extensive literature search was undertaken to identify all known and putative chromosomal variants that lead to AMR in *P. aeruginosa.* Among the 733 identified chromosomal AMR variants, 284 putative variants were classified as ‘natural variation’ as they were common in both antimicrobial-sensitive and AMR strains in our Global Dataset, and therefore deemed unlikely to contribute to an AMR phenotype. Most of these naturally occurring variants had been previously reported as putatively causing AMR, with little to no functional investigation. Importantly, no functionally validated AMR driving variants were re-classified as ‘natural variation’. Next, using mGWAS and comparative genomic analyses of the Global Dataset, we identified 75 previously unreported AMR variants associated with one or more AMR phenotypes (Table 2). In total, we identified 736 chromosomal variants in known AMR loci (Dataset 2), of which 374 were associated with AMR and 284 were natural variation.

**Table 1.** Summary of antimicrobial resistance prevalence across 10 clinically relevant antibiotics in the *Pseudomonas aeruginosa* Global Dataset.

| Antibiotic Class | Antibiotic | Res (%) | Int (%) | Sen (%) | No. isolates |
| --- | --- | --- | --- | --- | --- |
| Carbapenems | MEM | 682 (36) | 149 (8) | 682 (36) | 1875 |
|  | IPM | 186 (29) | 46 (7) | 419 (64) | 651 |
| Polymyxins | CST | 48 (5) | 0 (0) | 933 (95) | 981 |
| Fluoroquinolones | CIP | 664 (54) | 78 (6) | 477 (39) | 1219 |
| Cephalosporins | FEP | 157 (17) | 111(12) | 650 (71) | 918 |
|  | CAZ | 316 (27) | 121 (10) | 733 (63) | 1170 |
| Penicillins | TZP | 179 (24) | 52 (7) | 526 (69) | 757 |
|  | PIP | 185 (36) | 55 (11) | 272 (53) | 512 |
| Aminoglycosides | TOB | 294 (29) | 12 (1) | 695 (69) | 1001 |
|  | AMK | 170 (12) | 56 (4) | 1199 (84) | 1425 |

Abbreviations: Res, resistant; Int, intermediate; Sen, sensitive; MEM, meropenem; IPM, imipenem;
CST, colistin; CIP, ciprofloxacin; FEP, cefepime; CAZ, ceftazidime, TZP, piperacillin/tazobactam; PIP,
piperacillin; TOB, tobramycin; AMK, amikacin

**Table 2.**
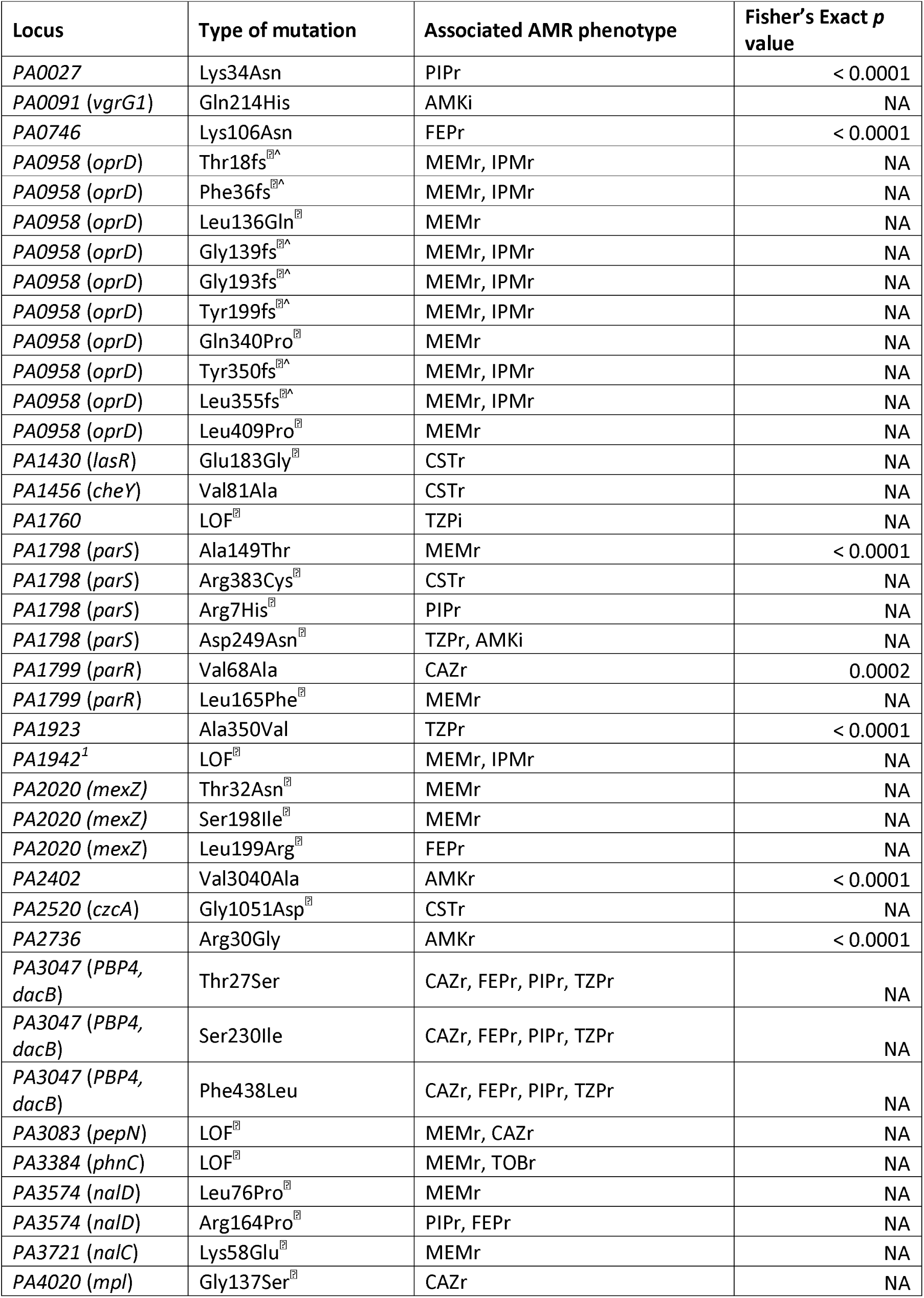
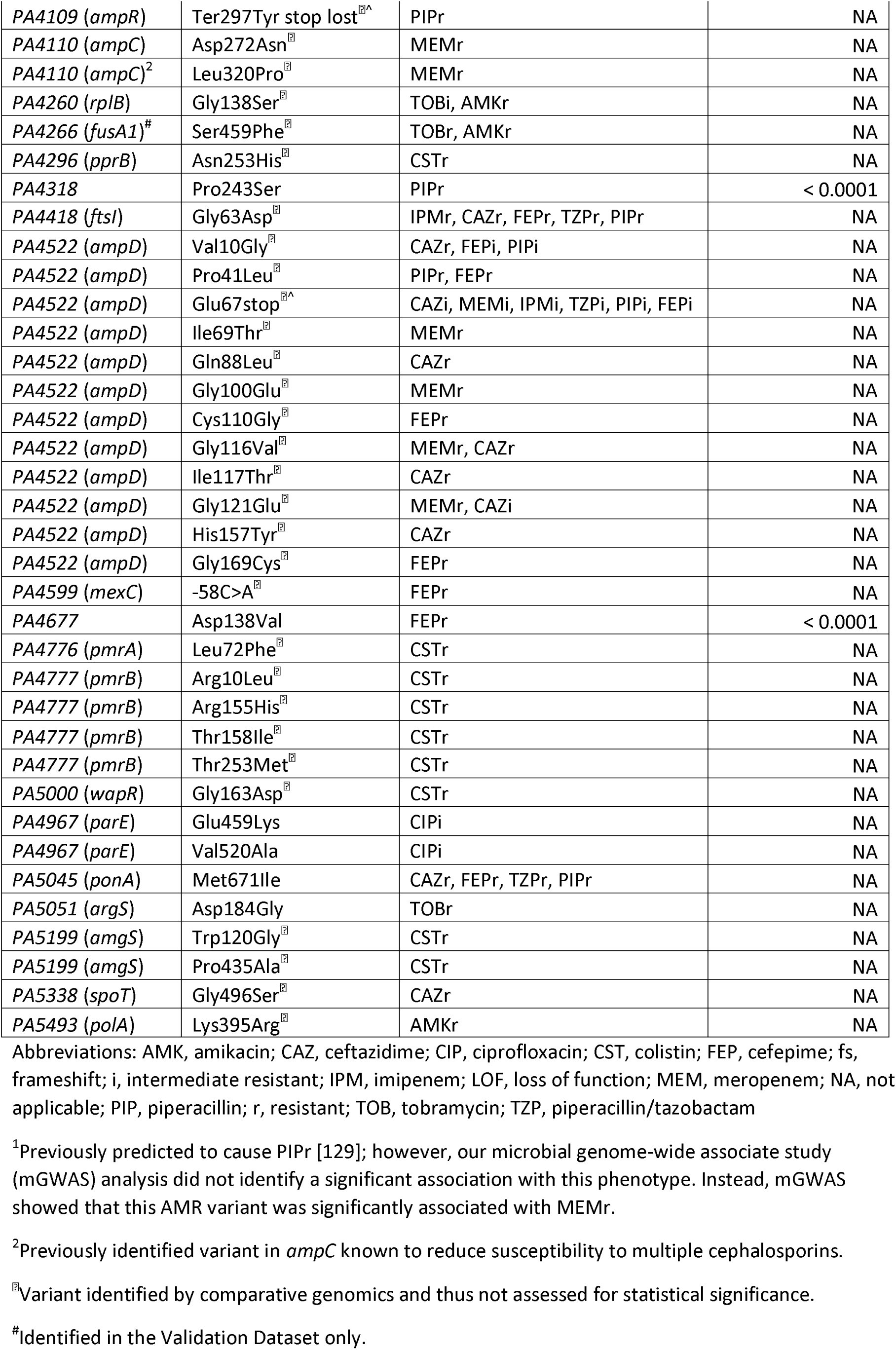
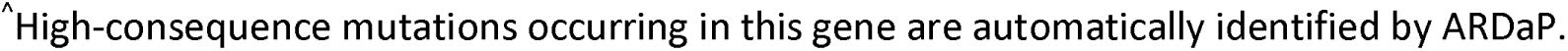
Novel antimicrobial resistance (AMR) variants identified in *Pseudomonas aeruginosa* by microbial genome-wide association study (mGWAS) or comparative genomic^0^ analyses.

### Predictive performance across Global Dataset

Although superior to CARD, abritAMR, AMRFinderPlus, and ResFinder still showed relatively poor bACCs for most antibiotics, with average bACCs of between 56 and 60%, well below ARDaP’s average bACC of 85% (Figure 1A).

**Figure 1.**
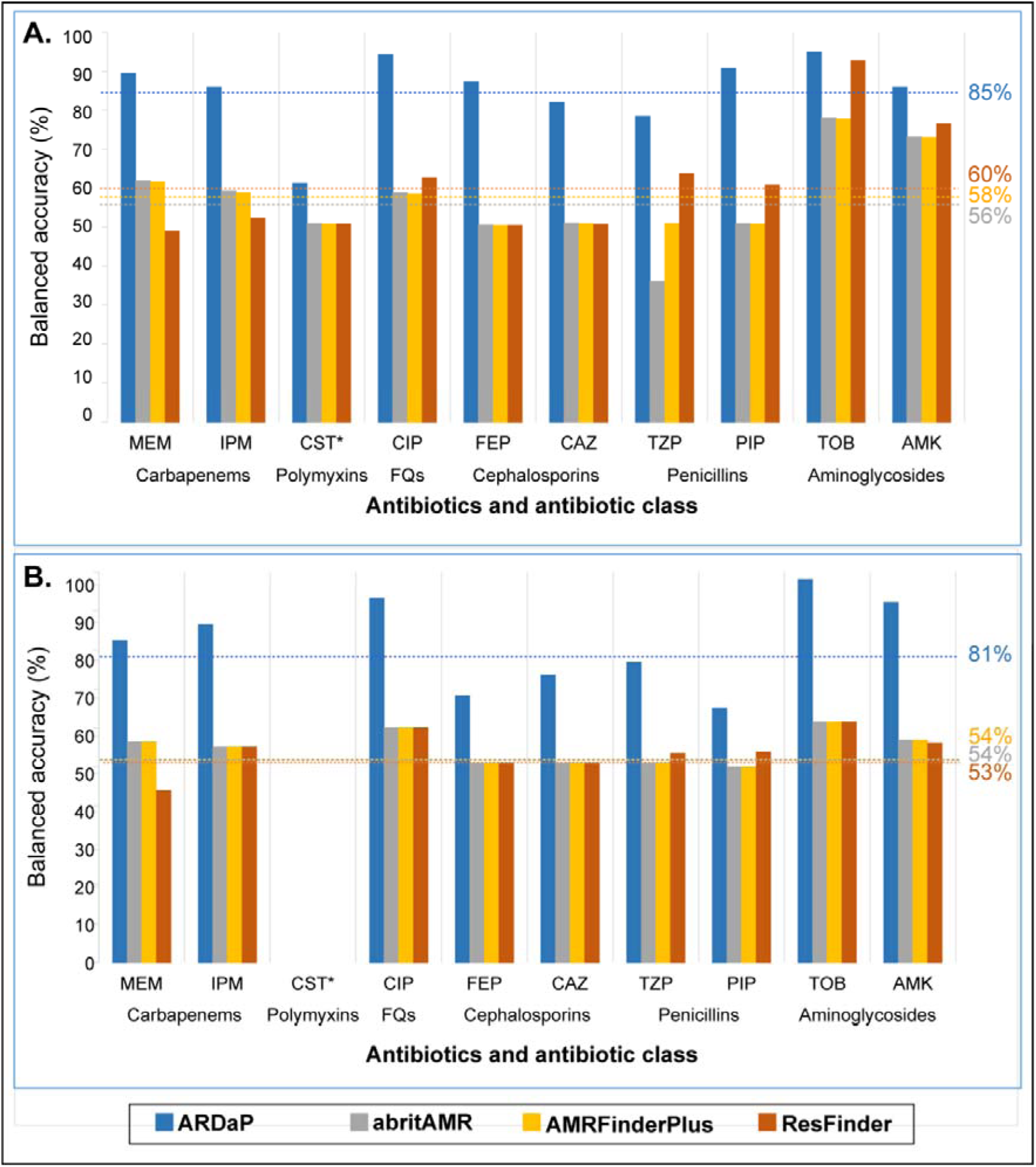
Balanced accuracy of ARDaP, abritAMR, AMRFinderPlus, and ResFinder for antimicrobial resistance (AMR) prediction in *Pseudomonas aeruginosa*. Software comparisons across ten clinically relevant antibiotics were undertaken against (A) the Global Dataset (*n*=1877 isolates) and (B) the Validation Dataset (*n*=102 isolates). For both datasets, and for all 10 antibiotics, ARDaP outperformed abritAMR, AMRFinderPlus, and ResFinder. To enable comparison with existing AMR prediction software, isolates with intermediate AMR were removed prior to analysis. Abbreviations: AMK, amikacin; CAZ, ceftazidime; CIP, ciprofloxacin; CST, colistin; FEP, cefepime; FQs, fluoroquinolones; IPM, imipenem; MEM, meropenem; PIP, piperacillin; TZP, piperacillin/tazobactam; TOB, tobramycin. *No strains in the Validation Dataset exhibited CST AMR; as such, balanced accuracy could not be calculated for this antibiotic.

The best abritAMR, AMRFinderPlus, and ResFinder predictions were achieved for the aminoglycosides, with average bACCs of 75%, 75%, and 82%, respectively, although these rates were lower than ARDaP’s average aminoglycoside bACC of 92%. abritAMR, AMRFinderPlus, and ResFinder AMR prediction for all other antibiotics showed poor to very poor bACCs. AMRFinderPlus had a bACC of just 50% for the penicillins, cephalosporins, and colistin – the same predictive capacity as a coin flip – and had only a slightly better bACC for ciprofloxacin (58%), imipenem (58%), and meropenem (61%).

ResFinder also had a bACC of just 50% for cephalosporins and colistin, and performed worse for carbapenems (average bACC of 50%) than AMRFinderPlus, although it was better for penicillins (average bACC of 62%) and ciprofloxacin (bACC of 62%). abritAMR was the worst at predicting penicillin phenotypes, with an average bACC of just 43%, worse than a coin flip; its performance was otherwise identical to AMRFinderPlus. In contrast, ARDaP surpassed abritAMR, AMRFinderPlus, and ResFinder across all 10 antibiotics, with bACCs ranging from 60% (colistin) to 94% (tobramycin) (Figure 1A).

### Predictive performance across Validation Dataset

We next tested abritAMR, AMRFinderPlus, ARDaP, and ResFinder across the Validation Dataset of 102 Australian clinical *P. aeruginosa* strains (Table S2) to determine each software’s performance in an analysis-naïve dataset. As no strains in the Validation Dataset displayed colistin AMR, the bACC for this antibiotic could not be assessed. These strains otherwise exhibited similar AMR rates to the Global Dataset, ranging from 26% for meropenem to 58% for piperacillin (Table S4).

Overall, ARDaP had high predictive accuracy (average bACC of 81%) across all antibiotics (Figure 1B), vastly outperforming ResFinder (average bACC of 53%), abritAMR, and AMRFinderPlus (average bACC of 54% each). Notably, the ResFinder meropenem bACC, at just 43%, yielded worse performance than a coin flip.

### Inclusion of the novel AMR variants identified in the Global Dataset

The inclusion of these markers increased Validation Dataset sensitivity by an average of 4% (range 0 to 27%) depending on antibiotic, with the sensitivity of most antibiotics (meropenem, imipenem, ciprofloxacin, cefepime, and piperacillin/tazobactam) remaining unchanged. Amikacin increased the most (27%) due to the inclusion of a SNP in *rplB* (Gly138Ser), followed by tobramycin at 4%, and piperacillin and ceftazidime at 3% each.”

### ARDaP performance between the Global and Validation Datasets

Whilst ARDaP bACC between the datasets were broadly similar, there was a greater proportion of false-positive and false-negative variants encoding AMR towards piperacillin (32% difference), tobramycin (24% difference), cefepime (19% difference), amikacin (12% difference), and meropenem (9% difference) in the Validation Dataset. In contrast, there was a greater proportion of false-positive and false-negative variants encoding amikacin AMR (5% difference) in the Global Dataset (Figure 1).

Comparative genomic analysis of Validation Dataset isolates that yielded false-negative aminoglycoside AMR predictions identified that many belonged to a single multilocus sequence type (ST), ST801, also known as AUST-06. Among 23/24 aminoglycoside-AMR ST801 isolates, a clade-specific missense variant in elongation factor G (FusA1 S459F) was identified; this SNP was not observed in other Global or Validation Dataset isolates. The remaining aminoglycoside-AMR ST801 strain, SCHI0010.S.1, encoded *AAC*(6’)-IIa, an aminoglycoside-modifying enzyme. Inclusion of FusA1 S459F into our AMR database significantly increased ARDaP bACCs for the Validation Dataset by an average 19% for both amikacin and tobramycin, raising them to 95% and 90%, respectively, with no impact on Global Dataset bACC.

### Precision and recall among AMR software

ARDaP demonstrated excellent precision and recall for predicting antimicrobial-sensitive and AMR phenotypes, ranging from 73% (average AMR recall) to 96% (average sensitivity recall) (Figure 2A). In contrast, abritAMR ranged from 54% (average AMR precision) to 62% (average sensitivity precision) (Figure 2B), AMRFinderPlus ranged from 54% (average AMR precision) to 62% (average AMR recall) (Figure 2C), and ResFinder ranged from only 42% (average AMR precision) to 68% (average sensitivity precision) (Figure 2D).

**Figure 2.**
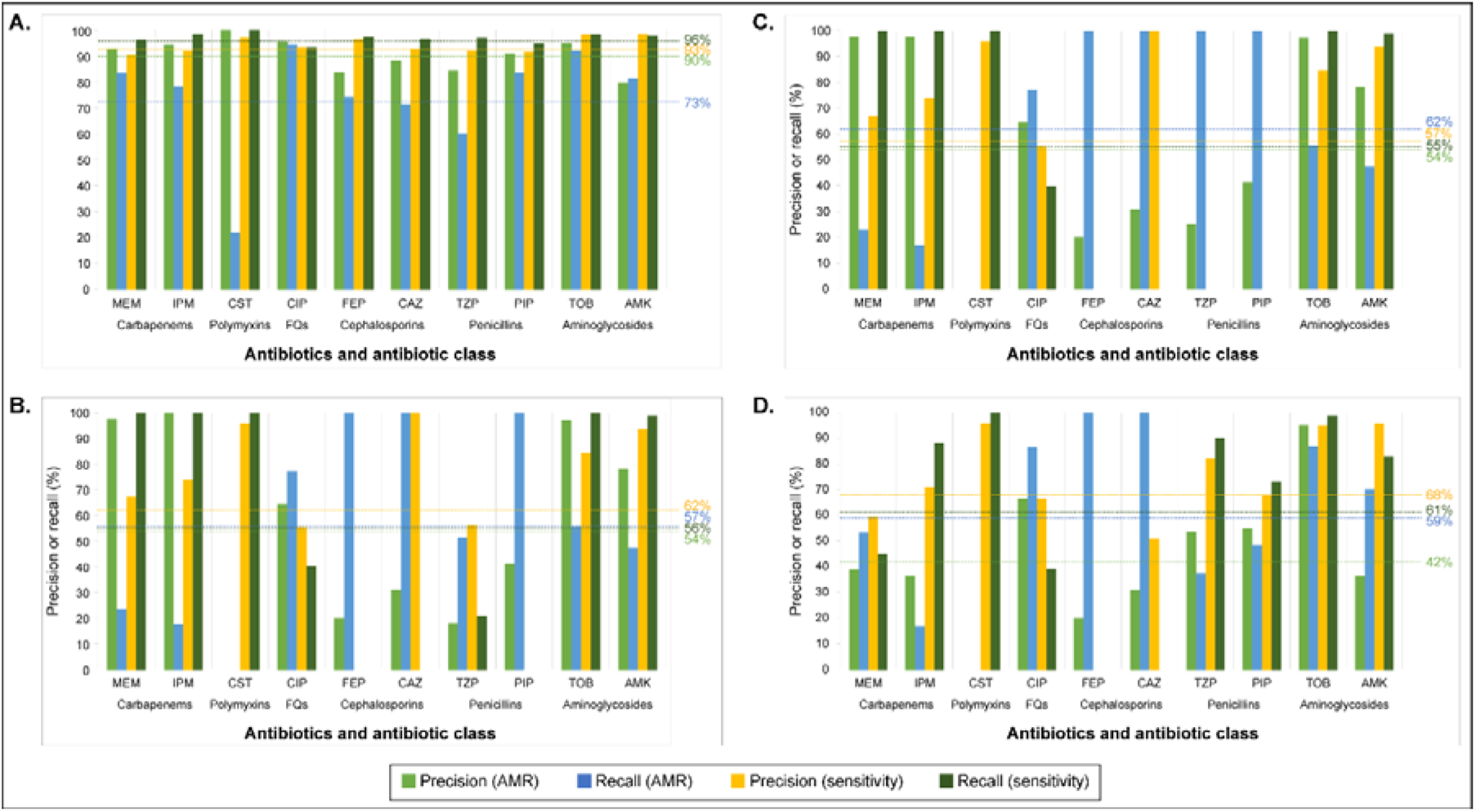
Precision and recall of ARDaP, abritAMR, AMRFinderPlus, and ResFinder across the Global Dataset (*n*=1877 strains). Precision and recall metrics for both antimicrobial-sensitive and antimicrobial-resistant strains were highest using ARDaP (A; range 73-96%) with abritAMR (B; range 54-62%), AMRFinderPlus (B; range 55-62%) and ResFinder (C; range 42-68%) all performing worse in comparison. To enable comparison with existing AMR prediction software, isolates with intermediate AMR were removed prior to analysis. Abbreviations: AMK, amikacin; CAZ, ceftazidime; CIP, ciprofloxacin; CST, colistin; FEP, cefepime; FQs, fluoroquinolones; IPM, imipenem; MEM, meropenem; PIP, piperacillin; TZP, piperacillin/tazobactam; TOB, tobramycin. N.B. Precision (resistance) is also known as positive predictive value (PPV) and precision (sensitivity) is also known as negative predictive value (NPV).

For colistin, abritAMR, AMRFinderPlus, and ResFinder all yielded AMR precision and recall values of 0%; in other words, none of these tools identified colistin AMR variants in strains exhibiting a colistin AMR phenotype. In comparison, ARDaP identified colistin AMR strains with 21% recall and 100% precision. Similarly, abritAMR, AMRFinderPlus, and ResFinder all failed to predict cefepime sensitivity in any cefepime-sensitive strain (Figures 2B-2D), instead erroneously classing every strain as cefepime-resistant, whereas ARDaP correctly identified cefepime-sensitive strains with 96% precision and 96% recall (Figure 2A). All three tools also failed to predict ceftazidime sensitivity. In addition, abritAMR and AMRFinderPlus failed to identify piperacillin sensitivity in any of the tested strains, and AMRFinderPlus yielded failed to identify piperacillin/tazobactam sensitivity (Figures 2B and 2C).

### Predictive performance for intermediate resistance

The inclusion of isolates with intermediate resistance reduced ARDaP bACCs by between 1 to 13% (6% average), depending on the antibiotic (Table S5). Intermediate resistance inclusion was most detrimental to cefepime AMR prediction (-13%), followed by imipenem (-8%), meropenem, ciprofloxacin and ceftazidime (-7%), piperacillin/tazobactam and amikacin (-5%), piperacillin (-4%) and tobramycin (-1%). Colistin prediction was unchanged as no intermediate category exists for this antibiotic.

## Discussion

The increasing role of high-throughput sequencing in the clinic has driven the concomitant development of bioinformatic tools for AMR variant detection and antimicrobial phenotype prediction [115]. However, current gold standard AMR tools are limited in their accuracy and performance due to their heavy focus on AMR gene gain rather than AMR-conferring chromosomal variants, and their inability to detect the gamut of genetic mutations that can confer AMR (i.e. gene loss, indels, CNVs, and structural variants) [14]. In addition, most tools have primarily focused on detection of AMR genes rather than AMR phenotype prediction. These shortcomings become acutely evident when attempting to predict AMR in pathogens with complex resistomes like *P. aeruginosa* [8].

To address this issue, we first constructed a comprehensive and accurate database of AMR variants encoded by *P. aeruginosa*. Our database of 733 chromosomal AMR-conferring variants (Dataset 2) comprises 374 previously identified AMR variants that we confirmed were significantly associated with an AMR phenotype, along with 75 AMR variants identified in this study. A further 284 AMR variants identified by us, and others, were classed as natural variants due to their non-significant association with AMR strains (Table S3). These variants were included for two reasons: i) to allow ARDaP’s coverage algorithm to scan known AMR genes for novel, high-consequence mutations (e.g. frameshift mutations in *oprD* that lead to carbapenem AMR) while avoiding natural variation that does not impact function, ii) to substantially reduce the legwork involved in identifying putative novel AMR variants using mGWAS or comparative genomics, and iii) to reduce the over-reporting of natural variants as putatively AMR conferring. Our natural variant list also provides a valuable resource for minimising erroneous AMR variant reporting in future *P. aeruginosa* AMR variant discovery studies. Next, performance assessment of our ARDaP-compatible AMR database against the Global and Validation Datasets showed that our tool outstripped the predictive performance of current ‘gold standard’ AMR software across all 10 antibiotics, yielding average bACC of 85%, vs. just 56%, 58% and 60% for abriTAMR, AMRFinderPlus and ResFinder, respectively (Figure 1). This performance difference is due to our chromosomal AMR variant database, which directly links genotypes with individual antibiotic phenotypes, and provides weighted scores on variant contribution towards AMR development (Dataset 1). Our findings concur with a recent study of 654 *P. aeruginosa* genomes, which also found that CARD and ResFinder exhibited poor AMR prediction performance metrics across all 11 tested anti-pseudomonal antibiotics [8].

Although still inferior to ARDaP, abritAMR, AMRFinderPlus, and ResFinder performed best when predicting aminoglycoside phenotypes in the Global Dataset (Figure 1A), which is heavily populated with American and European strains (Table S1). These tools performed substantially worse when tested against the Australian Validation Dataset (Figure 1B), with the average aminoglycoside bACC dropping by 16% for AMRFinderPlus and 26% for ResFinder. ARDaP’s bACC also initially dropped by 18% for the aminoglycosides. Upon closer inspection, we found that this performance reduction was predominantly due to false-negative calls among the ST801 isolates, a geographically restricted clone that has only been reported in people with CF in Qld, Australia [116]. Inclusion of one novel *fusA1* variant identified with comparative genomics in our AMR variant database restored ARDaP’s performance to bACC of 90% and 95% for amikacin and tobramycin, respectively. This performance difference across isolate datasets can be attributed to two phenomena. The first is the predominance of aminoglycoside-modifying enzymes in the Global (33%) but not Validation (8%) Datasets, reflecting potential major differences in the geographic prevalence of these enzymes that requires further exploration. The second is the enrichment of CF-derived isolates in the Validation Dataset, which comprise 86% of the aminoglycoside AMR strains (Figure S1). These isolates have largely developed aminoglycoside AMR via chromosomal mutation rather than aminoglycoside-modifying enzyme acquisition; as such, abritAMR, AMRFinderPlus, and ResFinder exhibited poor aminoglycoside AMR predictive capacity due to their limited chromosomal AMR variant databases. These performance differences highlight the need for including isolates from diverse sources, disease states, and locales to provide the most relevant AMR prediction software benchmarking comparisons. Our results suggest that abritAMR, AMRFinderPlus, and ResFinder are not useful for predicting aminoglycoside AMR from CF-derived *P. aeruginosa*, particularly in the Australian context, although this finding requires further exploration across larger, geographically diverse datasets.

Our findings revealed important weaknesses in abritAMR, AMRFinderPlus, and ResFinder when used for phenotype prediction. All three tools yielded bACCs of just 50% for cephalosporin prediction, abritAMR and AMRFinderPlus yielded a bACC of just 50% for piperacillin (Figures 1A and 1B), and abritAMR performed worse than a coin flip for predicting piperacillin/tazobactam phenotypes, with a bACC of just 35%. These cases of under-performance were largely attributed to sensitive isolates being predicted as AMR (Figure 2). The inferior performance of abritAMR and AMRFinderPlus over ResFinder was further exacerbated by their software design; for most anti-pseudomonal antibiotics, these tools only predict phenotypes to the antibiotic class level. To facilitate direct software comparisons, AMR identified for a given antibiotic class by abritAMR and AMRFinderPlus was extrapolated to all antibiotics within that class, which likely led to higher imprecision or error due to differences in within-class antibiotic spectrum of activity. For instance, the poor abritAMR piperacillin/tazobactam bACC may be attributed to this tool only reporting ‘β-lactamase’ presence; however, the impact of this β-lactamase on piperacillin/tazobactam efficacy is not explicitly reported due to insufficient granularity. Based on our and other’s [8] collective findings, we strongly discourage the use of abritAMR, AMRFinderPlus, or ResFinder for *in silico* cephalosporin AMR prediction in *P. aeruginosa* as none of these tools are currently capable of accurately differentiating sensitive from AMR strains for these antibiotics. Further, abritAMR and AMRFinderPlus should not be used to predict penicillin susceptibility phenotypes in *P. aeruginosa* due to their insufficient resolution.

Colistin prediction proved the most challenging of the 10 tested antibiotics, yielding bACCs of 50% with abriTAMR, AMRFinderPlus, and ResFinder, and 60% with ARDaP. ARDaP the only tool able to correctly predict some colistin AMR strains; the other three tools erroneously classified all *P. aeruginosa* strains as colistin-sensitive (Figures 2B and 2C). Accurate colistin prediction may have been hampered by the purported unreliability of gradient diffusion methods (e.g. disc diffusions, ETESTS) to accurately measure colistin breakpoints due to poor antibiotic diffusion and Mueller-Hinton agar manufacturer differences [117, 118]. Our study has identified a major gap in understanding the basis of colistin AMR, and underscores the need for much more work in this area, especially given the increasing use of inhaled colistin in treatment-refractory, multidrug-resistant *P. aeruginosa* infections [119, 120].

Loss-of-function mutations affecting the specialised porin, OprD, are the most common cause of carbapenem AMR in *P. aeruginosa*, particularly in clinical isolates [9]. Although *oprD* is notoriously hypervariable [121], ARDaP’s ability to accurately identify functional OprD loss accounted for its high carbapenem predictive accuracy (average bACC of 89% vs. just 61% for AMRFinderPlus and 52% for ResFinder). This outcome highlights the complex nature of the *P. aeruginosa* resistome, the necessity of AMR variant curation efforts, the value of AMR prediction tools that can accurately detect the spectrum of chromosomal variants, and the value of species-specific AMR variant databases to achieve the most accurate AMR predictions.

Mutations leading to chromosomal cephalosporinase (*ampC*) overexpression are an important cause of β-lactam AMR [15, 122]. To predict *ampC* upregulation, our database includes function-altering mutations in genes known to directly or indirectly regulate *ampC*. These chromosomally-encoded variants, alongside acquired cephalosporinases, accounted for the high average ARDaP bACC observed for the cephalosporins (84% vs. just 50% with abritAMR, AMRFinderPlus, and ResFinder; Figure 1). The prominence of *ampC* over-expression-associated determinants provides further support that this mechanism is a major cause of acquired cephalosporin AMR, particularly in clinical *P. aeruginosa* isolates [122, 123]. In support of this hypothesis, Khaledi and colleagues demonstrated a considerably higher bACC for ceftazidime AMR prediction when using both transcriptomic and genomic data (82%) compared with just genomic data (67%) [9]. Genomics alone cannot currently identify all instances of *ampC* over-expression, either because up-regulation is the result of an epigenic change, or the mutation remains cryptic due to an incomplete understanding of *ampC* regulatory mechanisms. Indeed, a review of intrinsic β-lactamases by Juan and colleagues details the complexity of *ampC* expression and its intricate regulation, along with the challenge of corresponding elevated β-lactamase MICs driven by *ampC* upregulation to clinical AMR breakpoints [124]. Using a combination of genomic and transcriptomic data will likely lead to further improvements in AMR prediction for most antibiotics [9, 122].

Predicting intermediate resistance is exceedingly difficult from genomic data alone, even with complex machine learning algorithms that combine transcriptomic and genomic data [9]. We also encountered difficulties in predicting intermediate resistance, with the inclusion of intermediate strains dropping bACCs by up to 13% (Table S5). Possible explanations include the need to understand the contribution of stepwise variants in conferring decreased antibiotic susceptibility [15, 67, 125], subtle and rapidly reversible gene expression alterations [9] caused by methylation [126] or dynamic environmental stimuli, and undetected strain mixtures. Further refinement of our ARDaP-compatible database, such as capacity to analyse RNA-seq data, will continue to improve this critical yet understudied area. Nevertheless, the pioneering capacity of ARDaP to predict intermediate resistance, including stepwise mutations that lower the barrier to full AMR development, and to differentiate strain mixtures in metagenomic data, has important implications for detecting emerging AMR in *P. aeruginosa* and informing earlier treatment shifts [14].

Errors introduced during sample collection, metadata curation, specimen processing, or sequencing may be partially responsible for our inability to predict AMR with a 100% bACC for any antibiotic. For example, 11 strains in the Khaledi *et al*. dataset [9] possessed variants known to confer ceftazidime AMR (e.g. *bla_VIM-_*[2,4,47], *bla_OXA-2_*, *bla_GES-_*[1,5]) yet were reported as ceftazidime-sensitive, and 22 strains in the Kos *et al*. dataset were amikacin-sensitive, yet possessed the aminoglycoside-modifying enzyme gene *aac(6’)-Ib-cr*, known to cause amikacin AMR and reduced ciprofloxacin susceptibility [127]. Due to the presence of these known AMR variants, all tools identified these strains as AMR, contributing to imperfect bACC (Figure 1) and poor precision (Figure 2) for amikacin and ceftazidime. As we did not have access to these strains, it was not possible to retest their AMR phenotypes or to repeat genome sequencing; however, we hypothesise that these strains would generate different results upon phenotypic retesting or re-sequencing. Alternatively, these AMR variants may be present but functionally or transcriptionally inactive, resulting in false-positive predictions for these isolates that must be factored into future AMR prediction estimates.

We recognise several study limitations. First, some false-positive predictions were identified across all antibiotic classes with our ARDaP-compatible database; however, these rates were significantly lower than those reported by other software (Figure 1). Whilst not ideal, we chose to retain a small number of AMR variants that result in low-frequency false-positive predictions as: i) some strains may have reverted to a sensitive phenotype, despite encoding a known AMR variant; and ii) we included phenotypic data generated by others, which may harbour inaccuracies. Functional profiling will be essential to fully understand the contribution of each of these variants in conferring AMR. Second, although we aimed for a phylogenetically diverse Validation Dataset (Figure S1, S2), only isolates from Queensland, Australia, were included in this dataset, limiting geographic and genetic representation. Despite this shortcoming, the Validation Dataset proved extremely useful for identifying AMR variant database deficits across all three AMR tools, particularly those variants encoding AMR towards the aminoglycosides, piperacillin, cefepime, and meropenem, highlighting clear areas of need for future research efforts. Third, due to cost constraints, we only performed ciprofloxacin and meropenem ETESTs for the Validation Dataset isolates, with disc diffusions used for the remaining eight antibiotics, a less robust methodology that may have led to some minor discrepancies in antimicrobial phenotype assignments. Fourth, we did not test out ARDaP’s capacity to identify *P. aeruginosa* AMR variants from simulated and real metagenomic datasets and strain mixtures for this study as we have proven this capacity elsewhere [14, 106] nor were we able to compare software performance against ARESdb [128] due to the proprietary nature of this database. Finally, our study would have benefitted from the inclusion of transcriptomic data [9] to identify additional novel variants associated with AMR. Although our understanding of the molecular mechanisms of AMR in *P. aeruginosa* is improving rapidly in the genomics era, false-negative predictions still occur across all antibiotic classes. Using ARDaP, such false-negative strains can now be rapidly identified and targeted for future functional work to pinpoint novel AMR variants and mechanisms.

## Declarations

### Ethics approval and consent to participate

Clinical specimen collection was approved by The Prince Charles Hospital (HREC/13/QPCH/127 and HREC/2019/QPCH/48013) and Royal Brisbane and Women’s Hospital (DA: jl) Human Research Ethics Committees. Participants with CF or COPD provided written consent; a waiver of informed consent was approved for other specimens due to a low-to-negligible risk assessment.

### Consent for publication

Not applicable.

### Availability of data and materials

The datasets supporting the conclusions of this article are available in the NCBI GenBank (references NSXK00000000.1 and NSZK00000000.1) and Sequence Read Archive database via BioProject accession PRJNA761496 (Validation Dataset), and BioProject accessions PRJEB15036, PRJEB21341, PRJEB29539, PRJEB14771, PRJNA297679, PRJNA317143, PRJNA526797, PRJNA264310, PRJNA388357, PRJNA793523, PRJEB40140 and PRJNA532924 (Global Dataset). The *P. aeruginosa* AMR database constructed for this study is freely available at: https://github.com/dsarov/ARDaP/tree/master/Databases/Pseudomonas_aeruginosa_pao1. The ARDaP software is freely available at: https://github.com/dsarov/ARDaP. All code used in this manuscript is available at https://github.com/dsarov/P_aeruginosa_ARDaP_manuscript.

### Authors’ contributions

DEM drafted the initial manuscript, compiled the original *P. aeruginosa* AMR variant database, performed data analyses and laboratory work, and assisted with ARDaP software refinement; TB provided chronic obstructive pulmonary disease respiratory specimens that were used to retrieve *P. aeruginosa* isolates, assisted with obtaining ethics approvals for sputum and bronchial washing collection, and obtained study funding; SCB provided CF sputa that were used to retrieve *P. aeruginosa* isolates, and obtained ethics approval for sputum collection; KM provided bloodstream *P. aeruginosa* isolates, and obtained ethics approval for blood collection; EPP assisted with study conception, obtaining and overseeing ethics approvals, manuscript preparation, data analyses and laboratory work, provided student supervision, obtained study funding, and assisted with curation of the natural variant database; and DSS conceived and led the study, wrote the ARDaP software, refined the *P. aeruginosa* AMR variant database, assisted with manuscript preparation, data analyses and laboratory work, provided student supervision, and obtained study funding. All authors read and approved of the final manuscript.

## Supporting information

Combined supplementary figures

## Data Availability

The datasets supporting the conclusions of this article are available in the NCBI GenBank (references NSXK00000000.1 and NSZK00000000.1) and Sequence Read Archive database via BioProject accession PRJNA761496 (Validation Dataset), and BioProject accessions PRJEB15036, PRJEB21341, PRJEB29539, PRJEB14771, PRJNA297679, PRJNA317143, PRJNA526797, PRJNA264310, PRJNA388357, PRJNA793523, PRJEB40140 and PRJNA532924 (Global Dataset). The P. aeruginosa AMR database constructed for this study is freely available at: https://github.com/dsarov/ARDaP/tree/master/Databases/Pseudomonas_aeruginosa_pao1. The ARDaP software is freely available at: https://github.com/dsarov/ARDaP. All code used in this manuscript is available at https://github.com/dsarov/P_aeruginosa_ARDaP_manuscript.

## Acknowledgements

We wish to thank Jane Neill (Sunshine Coast Hospital and Health Service, Sunshine Coast University Hospital) for providing the chronic obstructive pulmonary samples used in this study, and Kay Ramsay (University of Queensland) and Tamieka Fraser (University of the Sunshine Coast) for laboratory support.

## References

1. Mancuso G, Midiri A, Gerace E, Biondo C: Bacterial antibiotic resistance: The most critical pathogens. Pathogens (Basel, Switzerland) 2021, 10(10):1310.

2. Bassetti M, Merelli M, Temperoni C, Astilean A: New antibiotics for bad bugs: where are we? Annals of Clinical Microbiology and Antimicrobials 2013, 12(1):22.

3. Alanis AJ: Resistance to antibiotics: are we in the post-antibiotic era? Archives of Medical Research 2005, 36(6):697–705.

4. Antimicrobial resistance: Tackling a crisis for the health and wealth of nations

5. G. B. D. Antimicrobial Resistance Collaborators: Global mortality associated with 33 bacterial pathogens in 2019: a systematic analysis for the Global Burden of Disease Study 2019. Lancet 2022, 400(10369):2221–2248.

6. Denissen J, Reyneke B, Waso-Reyneke M, Havenga B, Barnard T, Khan S, Khan W: Prevalence of ESKAPE pathogens in the environment: Antibiotic resistance status, community-acquired infection and risk to human health. Int J Hyg Environ Health 2022, 244:114006.

7. Haenni M, Hocquet D, Ponsin C, Cholley P, Guyeux C, Madec JY, Bertrand X: Population structure and antimicrobial susceptibility of *Pseudomonas aeruginosa* from animal infections in France. BMC Vet Res 2015, 11:9.

8. Mahfouz N, Ferreira I, Beisken S, von Haeseler A, Posch AE: Large-scale assessment of antimicrobial resistance marker databases for genetic phenotype prediction: a systematic review. Journal of Antimicrobial Chemotherapy 2020, 75(11):3099–3108.

9. Khaledi A, Weimann A, Schniederjans M, Asgari E, Kuo TH, Oliver A, Cabot G, Kola A, Gastmeier P, Hogardt M et al: Predicting antimicrobial resistance in *Pseudomonas aeruginosa* with machine learning-enabled molecular diagnostics. EMBO Mol Med 2020, 12(3):e10264.

10. Bortolaia V, Kaas RS, Ruppe E, Roberts MC, Schwarz S, Cattoir V, Philippon A, Allesoe RL, Rebelo AR, Florensa AF et al: ResFinder 4.0 for predictions of phenotypes from genotypes. Journal of Antimicrobial Chemotherapy 2020, 75(12):3491–3500.

11. Alcock BP, Raphenya AR, Lau TTY, Tsang KK, Bouchard M, Edalatmand A, Huynh W, Nguyen A- LV, Cheng AA, Liu S et al: CARD 2020: antibiotic resistome surveillance with the comprehensive antibiotic resistance database. Nucleic Acids Research 2019, 48(D1):D517–D525.

12. Hunt M, Mather AE, Sanchez-Buso L, Page AJ, Parkhill J, Keane JA, Harris SR: ARIBA: Rapid antimicrobial resistance genotyping directly from sequencing reads. Microbial genomics 2017, 3(10):e000131.

13. Zankari E, Allesoe R, Joensen KG, Cavaco LM, Lund O, Aarestrup FM: PointFinder: A novel web tool for WGS-based detection of antimicrobial resistance associated with chromosomal point mutations in bacterial pathogens. Journal of Antimicrobial Chemotherapy 2017, 72(10):2764–2768.

14. Madden DE, Webb JR, Steinig EJ, Currie BJ, Price EP, Sarovich DS: Taking the next-gen step: Comprehensive antimicrobial resistance detection from *Burkholderia pseudomallei*. EBioMedicine 2021, 63:103152.

15. Lister PD, Wolter DJ, Hanson ND: Antibacterial-resistant *Pseudomonas aeruginosa:* Clinical impact and complex regulation of chromosomally encoded resistance mechanisms. Clinical Microbiology Reviews 2009, 22(4):582–610.

16. Pesesky MW, Hussain T, Wallace M, Patel S, Andleeb S, Burnham CD, Dantas G: Evaluation of machine learning and rules-based approaches for predicting antimicrobial resistance profiles in Gram-negative Bacilli from whole genome sequence data. Frontiers in microbiology 2016, 7:1887.

17. Gupta SK, Padmanabhan BR, Diene SM, Lopez-Rojas R, Kempf M, Landraud L, Rolain JM: ARG-ANNOT, a new bioinformatic tool to discover antibiotic resistance genes in bacterial genomes. Antimicrobial agents and chemotherapy 2014, 58(1):212–220.

18. Stover CK, Pham XQ, Erwin AL, Mizoguchi SD, Warrener P, Hickey MJ, Brinkman FS, Hufnagle WO, Kowalik DJ, Lagrou M et al: Complete genome sequence of *Pseudomonas aeruginosa* PAO1, an opportunistic pathogen. Nature 2000, 406(6799):959–964.

19. Lopez-Causape C, Cabot G, Del Barrio-Tofino E, Oliver A: The versatile mutational resistome of *Pseudomonas aeruginosa*. Frontiers in microbiology 2018, 9:685.

20. Kos VN, Deraspe M, McLaughlin RE, Whiteaker JD, Roy PH, Alm RA, Corbeil J, Gardner H: The resistome of *Pseudomonas aeruginosa* in relationship to phenotypic susceptibility. Antimicrobial agents and chemotherapy 2015, 59(1):427–436.

21. Poole K: *Pseudomonas aeruginosa:* Resistance to the max. Frontiers in microbiology 2011, 2:65–65.

22. Richardot C, Plesiat P, Fournier D, Monlezun L, Broutin I, Llanes C: Carbapenem resistance in cystic fibrosis strains of *Pseudomonas aeruginosa* as a result of amino acid substitutions in porin OprD. International journal of antimicrobial agents 2015, 45(5):529–532.

23. Tomás M, Doumith M, Warner M, Turton JF, Beceiro A, Bou G, Livermore DM, Woodford N: Efflux pumps, OprD porin, AmpC beta-lactamase, and multiresistance in *Pseudomonas aeruginosa* isolates from cystic fibrosis patients. Antimicrobial agents and chemotherapy 2010, 54(5):2219–2224.

24. Lister PD, Wolter DJ, Hanson ND: Antibacterial-resistant *Pseudomonas aeruginosa:* Clinical impact and complex regulation of chromosomally encoded resistance mechanisms. Clinical Microbiology Reviews 2009, 22(4):582–610.

25. Bortolaia V, Kaas RS, Ruppe E, Roberts MC, Schwarz S, Cattoir V, Philippon A, Allesoe RL, Rebelo AR, Florensa AF et al: ResFinder 4.0 for predictions of phenotypes from genotypes. J Antimicrob Chemother 2020, 75(12):3491–3500.

26. Heffernan AJ, Sime FB, Sarovich DS, Neely M, Guerra-Valero Y, Naicker S, Cottrell K, Harris P, Andrews KT, Ellwood D et al: Pharmacodynamic evaluation of plasma and epithelial lining fluid exposures of amikacin against *Pseudomonas aeruginosa* in a dynamic *in vitro* hollow-fiber infection model. Antimicrobial agents and chemotherapy 2020, 64(9).

27. Lau CH, Krahn T, Gilmour C, Mullen E, Poole K: AmgRS-mediated envelope stress-inducible expression of the *mexXY* multidrug efflux operon of *Pseudomonas aeruginosa*. MicrobiologyOpen 2015, 4(1):121–135.

28. Bolard A, Plésiat P, Jeannot K: Mutations in gene *fusA1* as a novel mechanism of aminoglycoside resistance in clinical strains of *Pseudomonas aeruginosa*. Antimicrobial agents and chemotherapy 2018, 62(2).

29. Wardell SJT, Rehman A, Martin LW, Winstanley C, Patrick WM, Lamont IL: A large-scale whole-genome comparison shows that experimental evolution in response to antibiotics predicts changes in naturally evolved clinical *Pseudomonas aeruginosa*. Antimicrobial agents and chemotherapy 2019.

30. López-Causapé C, Sommer LM, Cabot G, Rubio R, Ocampo-Sosa AA, Johansen HK, Figuerola J, Cantón R, Kidd TJ, Molin S et al: Evolution of the *Pseudomonas aeruginosa* mutational resistome in an international cystic fibrosis clone. Sci Rep 2017, 7(1):5555.

31. Lopez-Causape C, Rubio R, Cabot G, Oliver A: Evolution of the *Pseudomonas aeruginosa* aminoglycoside mutational resistome *in vitro* and in the cystic fibrosis setting. Antimicrobial agents and chemotherapy 2018, 62(4).

32. Sherrard LJ, Tai AS, Wee BA, Ramsay KA, Kidd TJ, Ben Zakour NL, Whiley DM, Beatson SA, Bell SC: Within-host whole genome analysis of an antibiotic resistant *Pseudomonas aeruginosa* strain sub-type in cystic fibrosis. PLoS One 2017, 12(3):e0172179.

33. Feng Y, Jonker MJ, Moustakas I, Brul S, Ter Kuile BH: Dynamics of mutations during development of resistance by *Pseudomonas aeruginosa* against five antibiotics. Antimicrobial agents and chemotherapy 2016, 60(7):4229–4236.

34. Khaledi A, Weimann A, Schniederjans M, Asgari E, Kuo T-H, Oliver A, Cabot G, Kola A, Gastmeier P, Hogardt M et al: Predicting antimicrobial resistance in *Pseudomonas aeruginosa* with machine learning-enabled molecular diagnostics. EMBO molecular medicine 2020, 12(3):e10264–e10264.

35. Sanz-Garcia F, Hernando-Amado S, Martinez JL: Mutational evolution of *Pseudomonas aeruginosa* resistance to ribosome-targeting antibiotics. Front Genet 2018, 9:451.

36. Schurek KN, Marr AK, Taylor PK, Wiegand I, Semenec L, Khaira BK, Hancock RE: Novel genetic determinants of low-level aminoglycoside resistance in *Pseudomonas aeruginosa*. Antimicrobial agents and chemotherapy 2008, 52(12):4213–4219.

37. Sun E, Gill EE, Falsafi R, Yeung A, Liu S, Hancock REW: Broad-spectrum adaptive antibiotic resistance associated with *Pseudomonas aeruginosa* mucin-dependent surfing motility. Antimicrobial agents and chemotherapy 2018, 62(9).

38. Dotsch A, Becker T, Pommerenke C, Magnowska Z, Jansch L, Haussler S: Genomewide identification of genetic determinants of antimicrobial drug resistance in *Pseudomonas aeruginosa*. Antimicrobial agents and chemotherapy 2009, 53(6):2522–2531.

39. Berrazeg M, Jeannot K, Ntsogo Enguene VY, Broutin I, Loeffert S, Fournier D, Plesiat P: Mutations in beta-Lactamase *ampC* increase resistance of *Pseudomonas aeruginosa* isolates to antipseudomonal cephalosporins. Antimicrobial agents and chemotherapy 2015, 59(10):6248–6255.

40. Colque CA, Albarracín Orio AG, Feliziani S, Marvig RL, Tobares AR, Johansen HK, Molin S, Smania AM: Hypermutator *Pseudomonas aeruginosa* exploits multiple genetic pathways to develop multidrug resistance during long-term infections in the airways of cystic fibrosis patients. Antimicrobial agents and chemotherapy 2020, 64(5).

41. Slater CL, Winogrodzki J, Fraile-Ribot PA, Oliver A, Khajehpour M, Mark BL: Adding insult to injury: Mechanistic basis for how AmpC mutations allow *Pseudomonas aeruginosa* to accelerate cephalosporin hydrolysis and evade avibactam. Antimicrobial agents and chemotherapy 2020, 64(9).

42. Fernandez-Esgueva M, Lopez-Calleja AI, Mulet X, Fraile-Ribot PA, Cabot G, Huarte R, Rezusta A, Oliver A: Characterization of AmpC beta-lactamase mutations of extensively drug-resistant *Pseudomonas aeruginosa* isolates that develop resistance to ceftolozane/tazobactam during therapy. Enferm Infecc Microbiol Clin (Engl Ed) 2020, 38(10):474–478.

43. Moya B, Dotsch A, Juan C, Blazquez J, Zamorano L, Haussler S, Oliver A: Beta-lactam resistance response triggered by inactivation of a nonessential penicillin-binding protein. PLoS Pathog 2009, 5(3):e1000353.

44. Zamorano L, Reeve TM, Deng L, Juan C, Moya B, Cabot G, Vocadlo DJ, Mark BL, Oliver A: NagZ inactivation prevents and reverts beta-lactam resistance, driven by AmpD and PBP 4 mutations, in *Pseudomonas aeruginosa*. Antimicrobial agents and chemotherapy 2010, 54(9):3557–3563.

45. Barbosa C, Gregg KS, Woods RJ: Variants in *ampD* and *dacB* lead to *in vivo* resistance evolution of *Pseudomonas aeruginosa* within the central nervous system. Journal of Antimicrobial Chemotherapy 2020, 75(11):3405–3408.

46. Langaee TY, Gagnon L, Huletsky A: Inactivation of the *ampD* gene in *Pseudomonas aeruginosa* leads to moderate-basal-level and hyperinducible AmpC beta-lactamase expression. Antimicrobial agents and chemotherapy 2000, 44(3):583–589.

47. Juan C, Macia MD, Gutierrez O, Vidal C, Perez JL, Oliver A: Molecular mechanisms of beta-lactam resistance mediated by AmpC hyperproduction in *Pseudomonas aeruginosa* clinical strains. Antimicrobial agents and chemotherapy 2005, 49(11):4733–4738.

48. Bagge N, Ciofu O, Hentzer M, Campbell JI, Givskov M, Hoiby N: Constitutive high expression of chromosomal beta-lactamase in *Pseudomonas aeruginosa* caused by a new insertion sequence (IS1669) located in *ampD*. Antimicrobial agents and chemotherapy 2002, 46(11):3406–3411.

49. Quale J, Bratu S, Gupta J, Landman D: Interplay of efflux system, *ampC*, and *oprD* expression in carbapenem resistance of *Pseudomonas aeruginosa* clinical isolates. Antimicrobial agents and chemotherapy 2006, 50(5):1633–1641.

50. Tsutsumi Y, Tomita H, Tanimoto K: Identification of novel genes responsible for overexpression of *ampC* in *Pseudomonas aeruginosa* PAO1. Antimicrobial agents and chemotherapy 2013, 57(12):5987–5993.

51. Kong KF, Aguila A, Schneper L, Mathee K: *Pseudomonas aeruginosa* β-lactamase induction requires two permeases, AmpG and AmpP. BMC microbiology 2010, 10:328.

52. Cabot G, Lopez-Causape C, Ocampo-Sosa AA, Sommer LM, Dominguez MA, Zamorano L, Juan C, Tubau F, Rodriguez C, Moya B et al: Deciphering the resistome of the widespread *Pseudomonas aeruginosa* sequence type 175 international high-risk clone through whole-genome sequencing. Antimicrob Agents Chemother 2016, 60(12):7415–7423.

53. Clark ST, Sinha U, Zhang Y, Wang PW, Donaldson SL, Coburn B, Waters VJ, Yau YCW, Tullis DE, Guttman DS et al: Penicillin-binding protein 3 is a common adaptive target among *Pseudomonas aeruginosa* isolates from adult cystic fibrosis patients treated with beta-lactams. International journal of antimicrobial agents 2019, 53(5):620–628.

54. Del Barrio-Tofiño E, López-Causapé C, Cabot G, Rivera A, Benito N, Segura C, Montero MM, Sorlí L, Tubau F, Gómez-Zorrilla S et al: Genomics and susceptibility profiles of extensively drug-resistant *Pseudomonas aeruginosa* isolates from Spain. Antimicrobial agents and chemotherapy 2017, 61(11).

55. Sherrard LJ, Wee BA, Duplancic C, Ramsay KA, Dave KA, Ballard E, Wainwright CE, Grimwood K, Sidjabat HE, Whiley DM et al: Emergence and impact of *oprD* mutations in *Pseudomonas aeruginosa* strains in cystic fibrosis. Journal of cystic fibrosis: official journal of the European Cystic Fibrosis Society 2021.

56. Livermore DM: Interplay of impermeability and chromosomal beta-lactamase activity in imipenem-resistant *Pseudomonas aeruginosa*. Antimicrobial agents and chemotherapy 1992, 36(9):2046–2048.

57. Ocampo-Sosa AA, Cabot G, Rodriguez C, Roman E, Tubau F, Macia MD, Moya B, Zamorano L, Suarez C, Pena C et al: Alterations of OprD in carbapenem-intermediate and -susceptible strains of *Pseudomonas aeruginosa* isolated from patients with bacteremia in a Spanish multicenter study. Antimicrobial agents and chemotherapy 2012, 56(4):1703–1713.

58. Fraile-Ribot PA, Mulet X, Cabot G, Del Barrio-Tofino E, Juan C, Perez JL, Oliver A: *In vivo* emergence of resistance to novel cephalosporin-beta-lactamase inhibitor combinations through the duplication of amino acid D149 from OXA-2 beta-lactamase (OXA-539) in sequence type 235 *Pseudomonas aeruginosa*. Antimicrobial agents and chemotherapy 2017, 61(9).

59. Cabot G, Bruchmann S, Mulet X, Zamorano L, Moyà B, Juan C, Haussler S, Oliver A: *Pseudomonas aeruginosa* ceftolozane-tazobactam resistance development requires multiple mutations leading to overexpression and structural modification of AmpC. Antimicrobial agents and chemotherapy 2014, 58(6):3091–3099.

60. Perron K, Caille O, Rossier C, Van Delden C, Dumas JL, Kohler T: CzcR-CzcS, a two-component system involved in heavy metal and carbapenem resistance in *Pseudomonas aeruginosa*. J Biol Chem 2004, 279(10):8761–8768.

61. Higgins PG, Fluit AC, Milatovic D, Verhoef J, Schmitz FJ: Mutations in GyrA, ParC, MexR and NfxB in clinical isolates of *Pseudomonas aeruginosa*. International journal of antimicrobial agents 2003, 21(5):409–413.

62. Lee JK, Lee YS, Park YK, Kim BS: Alterations in the GyrA and GyrB subunits of topoisomerase II and the ParC and ParE subunits of topoisomerase IV in ciprofloxacin-resistant clinical isolates of *Pseudomonas aeruginosa*. International journal of antimicrobial agents 2005, 25(4):290–295.

63. Bruchmann S, Dötsch A, Nouri B, Chaberny IF, Häussler S: Quantitative contributions of target alteration and decreased drug accumulation to *Pseudomonas aeruginosa* fluoroquinolone resistance. Antimicrobial agents and chemotherapy 2013, 57(3):1361–1368.

64. Jalal S, Wretlind B: Mechanisms of quinolone resistance in clinical strains of *Pseudomonas aeruginosa*. Microbial drug resistance (Larchmont, NY) 1998, 4(4):257–261.

65. Alyaseen SA, Piper KE, Rouse MS, Steckelberg JM, Patel R: Selection of cross-resistance following exposure of *Pseudomonas aeruginosa* clinical isolates to ciprofloxacin or cefepime. Antimicrobial agents and chemotherapy 2005, 49(6):2543–2545.

66. Akasaka T, Tanaka M, Yamaguchi A, Sato K: Type II topoisomerase mutations in fluoroquinolone-resistant clinical strains of *Pseudomonas aeruginosa* isolated in 1998 and 1999: role of target enzyme in mechanism of fluoroquinolone resistance. Antimicrobial agents and chemotherapy 2001, 45(8):2263–2268.

67. Rehman A, Jeukens J, Levesque RC, Lamont IL: Gene-gene interactions dictate ciprofloxacin resistance in *Pseudomonas aeruginosa* and facilitate prediction of resistance phenotype from genome sequence data. Antimicrobial agents and chemotherapy 2021, 65(7):e0269620.

68. Chilam J, Argimon S, Limas MT, Masim ML, Gayeta JM, Lagrada ML, Olorosa AM, Cohen V, Hernandez LT, Jeffrey B et al: Genomic surveillance of *Pseudomonas aeruginosa* in the Philippines, 2013-2014. Western Pac Surveill Response J 2021, 12(2):4–18.

69. Lee JY, Ko KS: Mutations and expression of PmrAB and PhoPQ related with colistin resistance in *Pseudomonas aeruginosa* clinical isolates. Diagn Microbiol Infect Dis 2014, 78(3):271–276.

70. Moskowitz SM, Ernst RK, Miller SI: PmrAB, a two-component regulatory system of *Pseudomonas aeruginosa* that modulates resistance to cationic antimicrobial peptides and addition of aminoarabinose to lipid A. J Bacteriol 2004, 186(2):575–579.

71. Abraham N, Kwon DH: A single amino acid substitution in PmrB is associated with polymyxin B resistance in clinical isolate of *Pseudomonas aeruginosa*. FEMS Microbiol Lett 2009, 298(2):249–254.

72. Moskowitz SM, Brannon MK, Dasgupta N, Pier M, Sgambati N, Miller AK, Selgrade SE, Miller SI, Denton M, Conway SP et al: PmrB mutations promote polymyxin resistance of *Pseudomonas aeruginosa* isolated from colistin-treated cystic fibrosis patients. Antimicrobial agents and chemotherapy 2012, 56(2):1019–1030.

73. Owusu-Anim D, Kwon DH: Differential role of two-component regulatory systems (*phoPQ* and *pmrAB*) in polymyxin B susceptibility of *Pseudomonas aeruginosa*. Advances in microbiology 2012, 2(1).

74. Choi MJ, Ko KS: Mutant prevention concentrations of colistin for *Acinetobacter baumannii, Pseudomonas aeruginosa* and *Klebsiella pneumoniae* clinical isolates. Journal of Antimicrobial Chemotherapy 2014, 69(1):275–277.

75. Saito K, Akama H, Yoshihara E, Nakae T: Mutations affecting DNA-binding activity of the MexR repressor of *mexR-mexA-mexB-oprM* operon expression. J Bacteriol 2003, 185(20):6195–6198.

76. Vaez H, Safaei HG, Faghri J: The emergence of multidrug-resistant clone ST664 *Pseudomonas aeruginosa* in a referral burn hospital, Isfahan, Iran. Burns Trauma 2017, 5:27.

77. Braz VS, Furlan JP, Fernandes AF, Stehling EG: Mutations in NalC induce MexAB-OprM overexpression resulting in high level of aztreonam resistance in environmental isolates of *Pseudomonas aeruginosa*. FEMS Microbiol Lett 2016, 363(16).

78. Suresh M, Nithya N, Jayasree PR, Vimal KP, Manish Kumar PR: Mutational analyses of regulatory genes, *mexR*, *nalC*, *nalD* and *mexZ* of *mexAB-oprM* and *mexXY* operons, in efflux pump hyperexpressing multidrug-resistant clinical isolates of *Pseudomonas aeruginosa*. World journal of microbiology & biotechnology 2018, 34(6):83.

79. Yan J, Estanbouli H, Liao C, Kim W, Monk JM, Rahman R, Kamboj M, Palsson BO, Qiu W, Xavier JB: Systems-level analysis of NalD mutation, a recurrent driver of rapid drug resistance in acute *Pseudomonas aeruginosa* infection. PLoS Comput Biol 2019, 15(12):e1007562.

80. Sobel ML, Neshat S, Poole K: Mutations in *PA2491* (*mexS*) promote MexT-dependent *mexEF-oprN* expression and multidrug resistance in a clinical strain of *Pseudomonas aeruginosa*. J Bacteriol 2005, 187(4):1246–1253.

81. Juarez P, Broutin I, Bordi C, Plésiat P, Llanes C: Constitutive activation of *mexT* by amino acid substitutions results in MexEF-OprN overproduction in clinical isolates of *Pseudomonas aeruginosa*. Antimicrobial agents and chemotherapy 2018, 62(5).

82. Llanes C, Köhler T, Patry I, Dehecq B, van Delden C, Plésiat P: Role of the MexEF-OprN efflux system in low-level resistance of *Pseudomonas aeruginosa* to ciprofloxacin. Antimicrobial agents and chemotherapy 2011, 55(12):5676–5684.

83. Guénard S, Muller C, Monlezun L, Benas P, Broutin I, Jeannot K, Plésiat P: Multiple mutations lead to MexXY-OprM-dependent aminoglycoside resistance in clinical strains of *Pseudomonas aeruginosa*. Antimicrobial agents and chemotherapy 2014, 58(1):221–228.

84. Chuanchuen R, Wannaprasat W, Ajariyakhajorn K, Schweizer HP: Role of the MexXY multidrug efflux pump in moderate aminoglycoside resistance in *Pseudomonas aeruginosa* isolates from *Pseudomonas* mastitis. Microbiology and immunology 2008, 52(8):392–398.

85. Hocquet D, Muller A, Blanc K, Plesiat P, Talon D, Monnet DL, Bertrand X: Relationship between antibiotic use and incidence of MexXY-OprM overproducers among clinical isolates of *Pseudomonas aeruginosa*. Antimicrobial agents and chemotherapy 2008, 52(3):1173–1175.

86. Islam S, Jalal S, Wretlind B: Expression of the MexXY efflux pump in amikacin-resistant isolates of *Pseudomonas aeruginosa*. Clinical microbiology and infection: the official publication of the European Society of Clinical Microbiology and Infectious Diseases 2004, 10(10):877–883.

87. Alvarez-Ortega C, Wiegand I, Olivares J, Hancock RE, Martinez JL: The intrinsic resistome of *Pseudomonas aeruginosa* to beta-lactams. Virulence 2011, 2(2):144–146.

88. Li XZ, Ma D, Livermore DM, Nikaido H: Role of efflux pump(s) in intrinsic resistance of *Pseudomonas aeruginosa*: active efflux as a contributing factor to beta-lactam resistance. Antimicrobial agents and chemotherapy 1994, 38(8):1742–1752.

89. Poole K, Heinrichs DE, Neshat S: Cloning and sequence analysis of an EnvCD homologue in *Pseudomonas aeruginosa*: regulation by iron and possible involvement in the secretion of the siderophore pyoverdine. Mol Microbiol 1993, 10(3):529–544.

90. Mine T, Morita Y, Kataoka A, Mizushima T, Tsuchiya T: Expression in *Escherichia coli* of a new multidrug efflux pump, MexXY, from *Pseudomonas aeruginosa*. Antimicrobial agents and chemotherapy 1999, 43(2):415–417.

91. Poole K, Gotoh N, Tsujimoto H, Zhao Q, Wada A, Yamasaki T, Neshat S, Yamagishi J, Li XZ, Nishino T: Overexpression of the *mexC-mexD-oprJ* efflux operon in *nfxB*-type multidrug-resistant strains of *Pseudomonas aeruginosa*. Mol Microbiol 1996, 21(4):713–724.

92. Köhler T, Michéa-Hamzehpour M, Henze U, Gotoh N, Curty LK, Pechère JC: Characterization of MexE-MexF-OprN, a positively regulated multidrug efflux system of *Pseudomonas aeruginosa*. Mol Microbiol 1997, 23(2):345–354.

93. Anuj SN, Whiley DM, Kidd TJ, Bell SC, Wainwright CE, Nissen MD, Sloots TP: Identification of *Pseudomonas aeruginosa* by a duplex real-time polymerase chain reaction assay targeting the *ecfX* and the *gyrB* genes. Diagn Microbiol Infect Dis 2009, 63(2):127–131.

94. Oliver A, Baquero F, Blázquez J: The mismatch repair system (*mutS*, *mutL* and *uvrD* genes) in *Pseudomonas aeruginosa*: molecular characterization of naturally occurring mutants. Mol Microbiol 2002, 43(6):1641–1650.

95. van Belkum A, Soriaga LB, LaFave MC, Akella S, Veyrieras JB, Barbu EM, Shortridge D, Blanc B, Hannum G, Zambardi G et al: Phylogenetic distribution of CRISPR-Cas systems in antibiotic-resistant *Pseudomonas aeruginosa*. mBio 2015, 6(6):e01796–01715.

96. Buhl M, Kästle C, Geyer A, Autenrieth IB, Peter S, Willmann M: Molecular evolution of extensively drug-resistant (XDR) *Pseudomonas aeruginosa* strains from patients and hospital environment in a prolonged outbreak. Frontiers in microbiology 2019, 10:1742.

97. Cabot G, López-Causapé C, Ocampo-Sosa AA, Sommer LM, Domínguez MA, Zamorano L, Juan C, Tubau F, Rodríguez C, Moyà B et al: Deciphering the resistome of the widespread *Pseudomonas aeruginosa* Sequence Type 175 international high-risk clone through whole-genome sequencing. Antimicrobial agents and chemotherapy 2016, 60(12):7415–7423.

98. CDC & FDA Antibiotic Resistance Isolate Bank for *Pseudomonas aeruginosa* [Available at: https://wwwn.cdc.gov/ARIsolateBank/Panel/PanelDetail?ID=12]

99. Ramanathan B, Jindal HM, Le CF, Gudimella R, Anwar A, Razali R, Poole-Johnson J, Manikam R, Sekaran SD: Next generation sequencing reveals the antibiotic resistant variants in the genome of *Pseudomonas aeruginosa*. PLoS One 2017, 12(8):e0182524.

100. Tsang KK, Maguire F, Zubyk HL, Chou S, Edalatmand A, Wright GD, Beiko RG, McArthur AG: Identifying novel β-lactamase substrate activity through *in silico* prediction of antimicrobial resistance. Microbial genomics 2021, 7(1).

101. Wardell SJT, Rehman A, Martin LW, Winstanley C, Patrick WM, Lamont IL: A large-scale whole-genome comparison shows that experimental evolution in response to antibiotics predicts changes in naturally evolved clinical *Pseudomonas aeruginosa*. Antimicrobial agents and chemotherapy 2019, 63(12).

102. Cortes-Lara S, Barrio-Tofiño ED, López-Causapé C, Oliver A, Gemara-Seimc Reipi Pseudomonas study Group: Predicting *Pseudomonas aeruginosa* susceptibility phenotypes from whole genome sequence resistome analysis. Clinical microbiology and infection: the official publication of the European Society of Clinical Microbiology and Infectious Diseases 2021, 27(11):1631–1637.

103. Sun Z, Yang F, Ji J, Cao W, Liu C, Ding B, Xu X: Dissecting the genotypic features of a fluoroquinolone-resistant Pseudomonas aeruginosa ST316 sublineage causing ear infections in Shanghai, China. Microbial genomics 2023, 9(4).

104. Su M, Satola SW, Read TD: Genome-based prediction of bacterial antibiotic resistance. Journal of clinical microbiology 2019, 57(3):e01405–01418.

105. Madden DE, McCarthy KL, Bell SC, Olagoke O, Baird T, Neill J, Ramsay KA, Kidd TJ, Stewart AG, Subedi S et al: Rapid fluoroquinolone resistance detection in *Pseudomonas aeruginosa* using mismatch amplification mutation assay-based real-time PCR. J Med Microbiol 2022, 71(10):001593.

106. Madden DE, Olagoke O, Baird T, Neill J, Ramsay KA, Fraser TA, Bell SC, Sarovich DS, Price EP: Express yourself: Quantitative real-time PCR assays for rapid chromosomal antimicrobial resistance detection in *Pseudomonas aeruginosa*. Antimicrobial agents and chemotherapy 2022, 66(5):e0020422.

107. Mobegi FM, Cremers AJ, de Jonge MI, Bentley SD, van Hijum SA, Zomer A: Deciphering the distance to antibiotic resistance for the pneumococcus using genome sequencing data. Sci Rep 2017, 7:42808.

108. Sarovich DS, Price EP: SPANDx: a genomics pipeline for comparative analysis of large haploid whole genome re-sequencing datasets. BMC research notes 2014, 7:618.

109. Crouch DJM, Bodmer WF: Polygenic inheritance, GWAS, polygenic risk scores, and the search for functional variants. Proc Natl Acad Sci U S A 2020, 117(32):18924–18933.

110. ARDaP - Antimicrobial Resistance Detection and Prediction. [https://github.com/dsarov/ARDaP]

111. Sherry NL, Horan KA, Ballard SA, Gonalves da Silva A, Gorrie CL, Schultz MB, Stevens K, Valcanis M, Sait ML, Stinear TP et al: An ISO-certified genomics workflow for identification and surveillance of antimicrobial resistance. Nat Commun 2023, 14(1):60.

112. Feldgarden M, Brover V, Haft DH, Prasad AB, Slotta DJ, Tolstoy I, Tyson GH, Zhao S, Hsu CH, McDermott PF et al: Validating the AMRFinder tool and resistance gene database by using antimicrobial resistance genotype-phenotype correlations in a collection of isolates. Antimicrobial agents and chemotherapy 2019, 63(11):e00483–00419.

113. Bekkar M, Djemaa HK, Alitouche TA: Evaluation measures for models assessment over imbalanced data sets. J Inf Eng Appl 2013, 3(10):27–38.

114. Hicks AL, Wheeler N, Sanchez-Buso L, Rakeman JL, Harris SR, Grad YH: Evaluation of parameters affecting performance and reliability of machine learning-based antibiotic susceptibility testing from whole genome sequencing data. PLoS Comput Biol 2019, 15(9):e1007349.

115. Van Goethem N, Descamps T, Devleesschauwer B, Roosens NHC, Boon NAM, Van Oyen H, Robert A: Status and potential of bacterial genomics for public health practice: a scoping review. Implement Sci 2019, 14(1):79.

116. Kidd TJ, Ritchie SR, Ramsay KA, Grimwood K, Bell SC, Rainey PB: *Pseudomonas aeruginosa* exhibits frequent recombination, but only a limited association between genotype and ecological setting. PLoS One 2012, 7(9):e44199.

117. Matuschek E, Ahman J, Webster C, Kahlmeter G: Antimicrobial susceptibility testing of colistin - evaluation of seven commercial MIC products against standard broth microdilution for *Escherichia coli*, *Klebsiella pneumoniae*, *Pseudomonas aeruginosa*, and *Acinetobacter* spp. Clinical microbiology and infection: the official publication of the European Society of Clinical Microbiology and Infectious Diseases 2018, 24(8):865–870.

118. Bassetti M, Vena A, Croxatto A, Righi E, Guery B: How to manage *Pseudomonas aeruginosa* infections. Drugs Context 2018, 7:212527.

119. Gurjar M: Colistin for lung infection: an update. J Intensive Care 2015, 3(1):3.

120. Sabuda DM, Laupland K, Pitout J, Dalton B, Rabin H, Louie T, Conly J: Utilization of colistin for treatment of multidrug-resistant *Pseudomonas aeruginosa*. Can J Infect Dis Med Microbiol 2008, 19(6):413–418.

121. Li H, Luo YF, Williams BJ, Blackwell TS, Xie CM: Structure and function of OprD protein in *Pseudomonas aeruginosa*: from antibiotic resistance to novel therapies. Int J Med Microbiol 2012, 302(2):63–68.

122. Cabot G, Ocampo-Sosa AA, Tubau F, Macia MD, Rodriguez C, Moya B, Zamorano L, Suarez C, Pena C, Martinez-Martinez L et al: Overexpression of *AmpC* and efflux pumps in *Pseudomonas aeruginosa* isolates from bloodstream infections: prevalence and impact on resistance in a Spanish multicenter study. Antimicrobial agents and chemotherapy 2011, 55(5):1906–1911.

123. Lee JY, Ko KS: OprD mutations and inactivation, expression of efflux pumps and AmpC, and metallo-β-lactamases in carbapenem-resistant *Pseudomonas aeruginosa* isolates from South Korea. International journal of antimicrobial agents 2012, 40(2):168–172.

124. Juan C, Torrens G, González-Nicolau M, Oliver A: Diversity and regulation of intrinsic beta-lactamases from non-fermenting and other Gram-negative opportunistic pathogens. FEMS Microbiol Rev 2017, 41(6):781–815.

125. Levy SB, Bonnie M: Antibacterial resistance worldwide: Causes, challenges and responses. Nature Medicine 2004, 10(12S):S122–S129.

126. Law COK, Huang C, Pan Q, Lee J, Hao Q, Chan T-F, Lo NWS, Ang IL, Koon A, Ip M et al: A small RNA transforms the multidrug resistance of *Pseudomonas aeruginosa* to drug susceptibility. Mol Ther Nucleic Acids 2019, 16:218–228.

127. Robicsek A, Strahilevitz J, Jacoby GA, Macielag M, Abbanat D, Park CH, Bush K, Hooper DC: Fluoroquinolone-modifying enzyme: a new adaptation of a common aminoglycoside acetyltransferase. Nat Med 2006, 12(1):83–88.

128. Ferreira I, Beisken S, Lueftinger L, Weinmaier T, Klein M, Bacher J, Patel R, von Haeseler A, Posch AE: Species Identification and Antibiotic Resistance Prediction by Analysis of Whole-Genome Sequence Data by Use of ARESdb: an Analysis of Isolates from the Unyvero Lower Respiratory Tract Infection Trial. Journal of clinical microbiology 2020, 58(7).

129. Dötsch A, Becker T, Pommerenke C, Magnowska Z, Jänsch L, Häussler S: Genomewide identification of genetic determinants of antimicrobial drug resistance in *Pseudomonas aeruginosa*. Antimicrobial agents and chemotherapy 2009, 53(6):2522–2531.

