## Supplementary material for "Keeping up with the pathogens: Improved antimicrobial resistance detection and prediction from *Pseudomonas* aeruginosa genomes": Combined supplementary figures

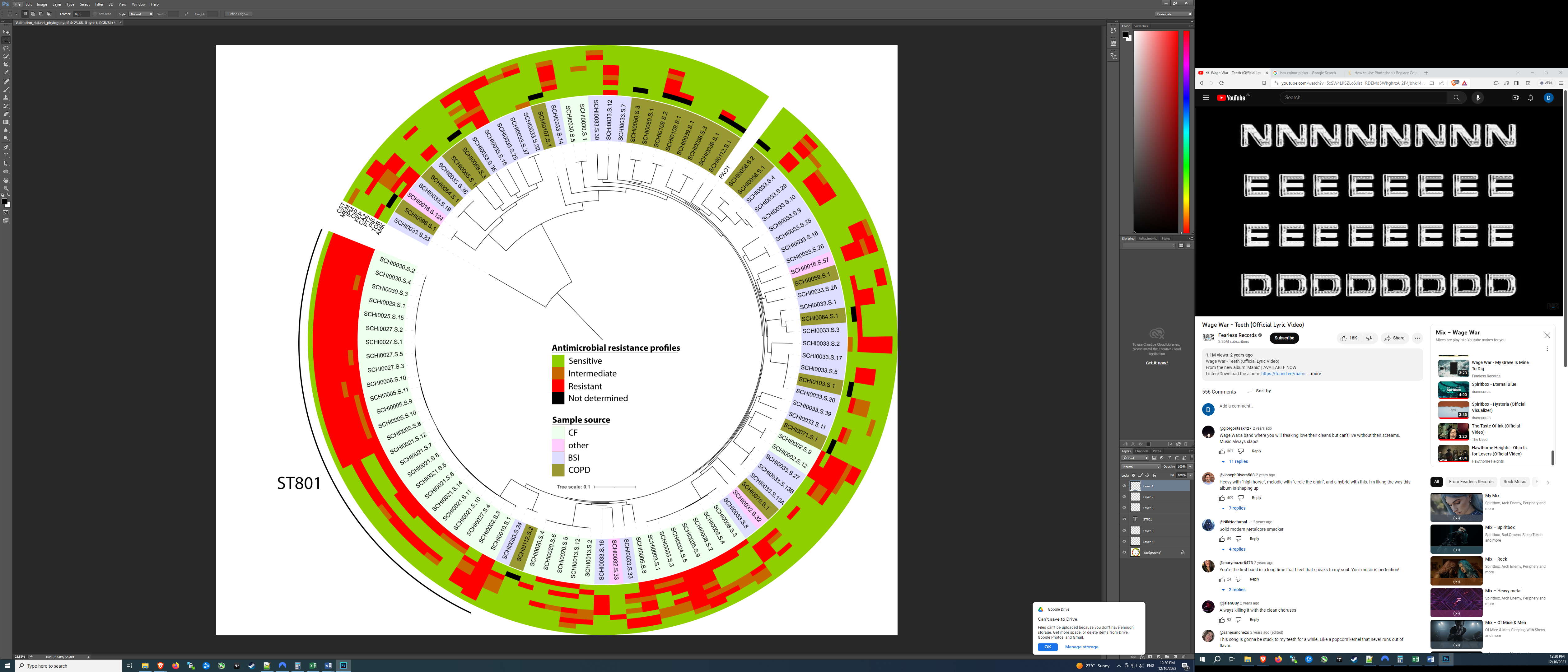

**Figure S1. Maximum likelihood phylogenomic analysis of the *Pseudomonas aeruginosa* ‘Validation Dataset’ generated in this study (*n*=102), along with their associated antimicrobial resistance profiles and disease origin.** *P. aeruginosa* PAO1 was used as the reference for alignment and single-nucleotide polymorphism identification. Variants were identified using the SPANDx v4.0.3 pipeline[1] with phylogenetic trees drawn with FastTree v2.1.10.[2] Shading represents isolate source and outer rings represent the antimicrobial resistance profile of the isolate. Abbreviations: AMK, amikacin; BSI, blood stream infection; CAZ, ceftazidime; CIP, ciprofloxacin; CST, colistin; CF, cystic fibrosis; COPD, chronic obstructive pulmonary disease; FEP, cefepime; IPM, imipenem; MEM, meropenem; PIP, piperacillin; ST, sequence type; TZP, piperacillin/tazobactam; TOB, tobramycin.

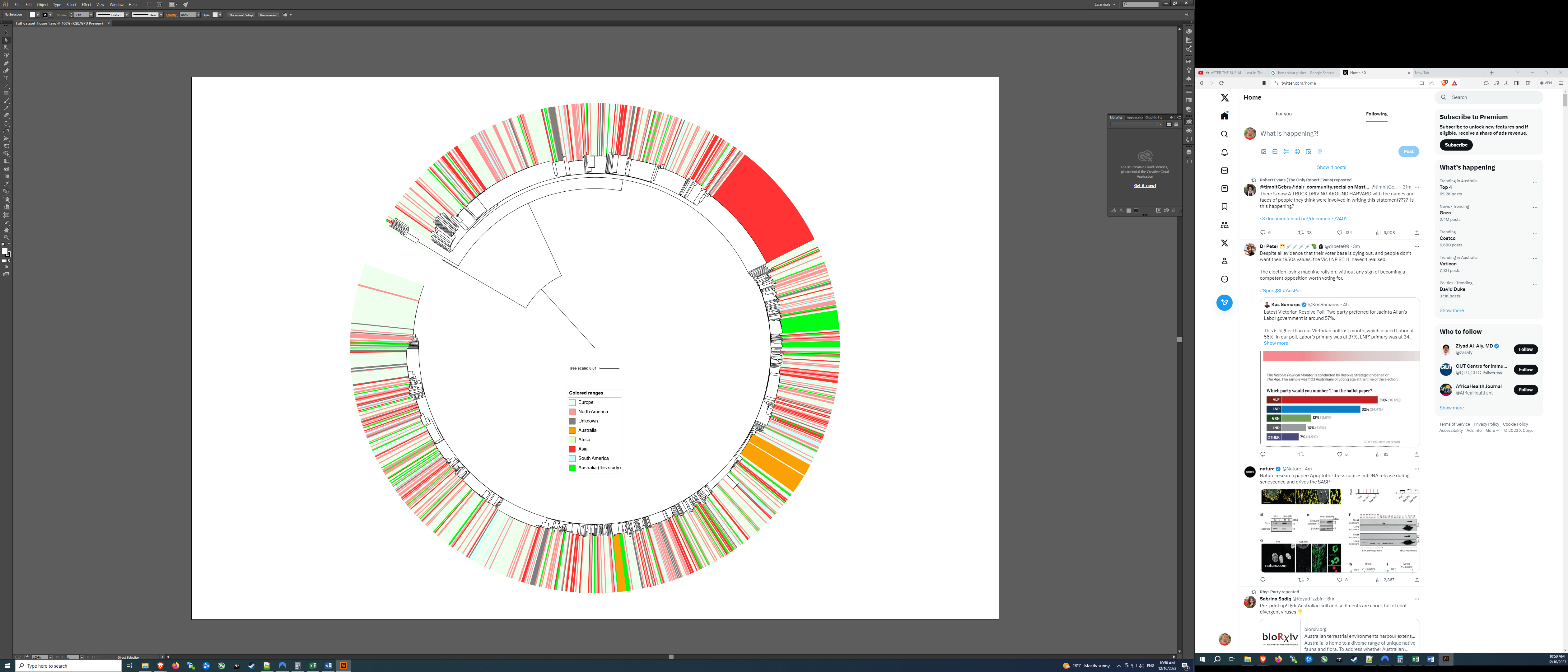

**Figure S2. Maximum likelihood phylogenomic analysis of all 1979 *Pseudomonas aeruginosa* strains examined in this study.** *P. aeruginosa* PAO1 was used as the reference genome for alignment and single-nucleotide polymorphism identification. Variants were identified using the SPANDx v4.0.3 pipeline[1], with phylogenetic trees drawn with FastTree v2.1.10[2]. Shading represents country of origin. The new Validation Dataset isolates generated in the current study (neon green) are distributed throughout the entire phylogeny, reflecting a panmictic population structure.

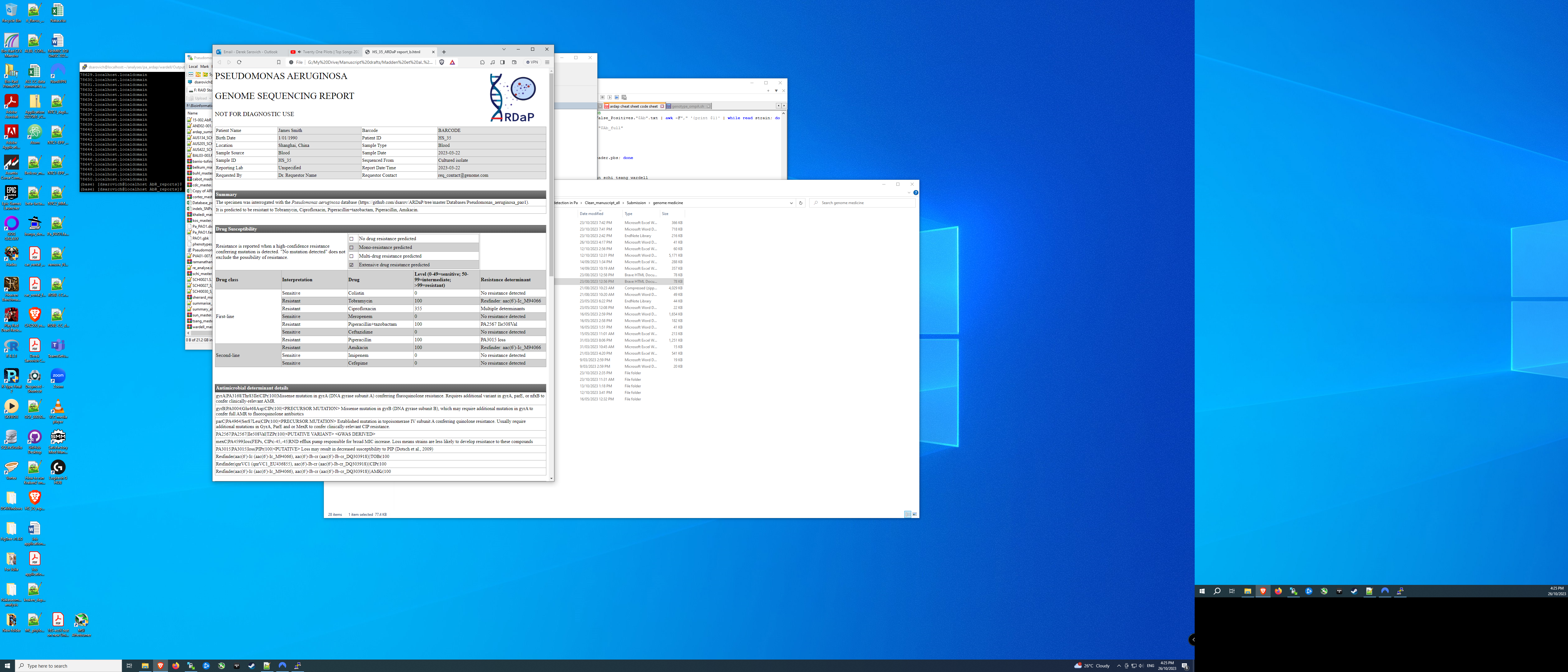

**Figure S3. Example clinician-friendly report produced by ARDaP for Chinese *Pseudomonas aeruginosa* strain HS_35.** This report summarises the predicted antimicrobial susceptibility profile, the individual drug susceptibility score, the drug susceptibility status (no resistance through to extensive drug resistance) for HS_35, and the corresponding antimicrobial resistance variants identified in this isolate.

**References**

1.	Sarovich DS, Price EP. SPANDx: a genomics pipeline for comparative analysis of large haploid whole genome re-sequencing datasets. BMC Res Notes **2014**; 7: 618.

2.	Price MN, Dehal PS, Arkin AP. FastTree 2 – approximately maximum-likelihood trees for large alignments. PLoS One **2010**; 5(3): e9490.
