## Supplementary material for "Keeping up with the pathogens: Improved antimicrobial resistance detection and prediction from *Pseudomonas* aeruginosa genomes": Table S1

**Table S1.** Summary of *Pseudomonas aeruginosa* Global Dataset strains with paired AMR and genomic data.

| **Study** | **No. strains^a^** | **Description** |
| --- | --- | --- |
| Kos *et al*., 2015 (1) | 385 | Diverse isolates collected from multiple geographical locations between 2003 and 2012 |
| van Belkum *et al*., 2015 (2) | 271^b^ | Diverse clinical isolates, mostly collected in Europe and USA, over a 25-year period |
| Cabot *et al*., 2016 (3) | 22 | MDR ST-175 isolates obtained from patients in 8 Spanish and 4 French hospitals, collected in 2008 and 2009 |
| Sherrard *et al*., 2017 (4) | 11 | MDR isolates collected from a cystic fibrosis patient in Queensland, Australia, between 2007 and 2014 |
| Ramanathan *et al*., 2017 (5) | 10 | Isolates collected from patients at a Malaysian hospital in 2009 and 2010 |
| Del Barrio-Tofiño *et al*., 2017 (6) | 41 | MDR clinical isolates collected in Spain, 2015 |
| CDC-FDA Collection, 2018 (7) | 54 | MDR isolates from diverse clinical sources that represent the diversity of AMR phenotypes |
| Buhl *et al*., 2019 (8) | 45 | Clinical multidrug-resistant isolates obtained from two German hospitals during a four-year outbreak |
| Wardell *et al*., 2019 (9) | 39 | PAO1-derived isolates subjected to *in vitro* evolution for AMR development |
| Khaledi *et al*., 2020 (10) | 411 | Genetically diverse isolates with MDR obtained from European clinical cases. Temporal range unknown |
| Tsang *et al*., 2021 (11) | 102 | MDR isolates collected from patients admitted to Canadian hospitals between 2015 and 2018 |
| Cortes-Lara *et al.,* 2021 (12) | 288 | Diverse collection of isolates from Spanish hospitals. Collected in 2017 |
| Sun et al., 2023 (13) | 197 | Characterization of the genotypic features of an emerging ST316 sub lineage causing ear infections in Shanghai |
| **Total** | **1,391** |  |

Abbreviations: AMR, antimicrobial-resistant; MDR, multidrug-resistant; ST, multi-locus sequence-type. ^a^Excludes genomes that were found to be of very low quality/coverage or missing from public databases. ^b^van Belkum *et al*., 2015 also included isolates from Kos *et al*., 2015. Strain numbers are representative of unique isolates only

**References**

1.	Kos VN, Déraspe M, McLaughlin RE, Whiteaker JD, Roy PH, Alm RA, Corbeil J, Gardner H. 2015. The resistome of *Pseudomonas aeruginosa* in relationship to phenotypic susceptibility. Antimicrob Agents Chemother 59:427-436.

2.	van Belkum A, Soriaga LB, LaFave MC, Akella S, Veyrieras J-B, Barbu EM, Shortridge D, Blanc B, Hannum G, Zambardi G, Miller K, Enright MC, Mugnier N, Brami D, Schicklin S, Felderman M, Schwartz AS, Richardson TH, Peterson TC, Hubby B, Cady KC. 2015. Phylogenetic distribution of CRISPR-Cas systems in antibiotic-resistant *Pseudomonas aeruginosa*. mBio 6:e01796-15.

3.	Cabot G, López-Causapé C, Ocampo-Sosa AA, Sommer LM, Domínguez MÁ, Zamorano L, Juan C, Tubau F, Rodríguez C, Moyà B, Peña C, Martínez-Martínez L, Plesiat P, Oliver A. 2016. Deciphering the resistome of the widespread *Pseudomonas aeruginosa* sequence type 175 international high-risk clone through whole-genome sequencing. Antimicrob Agents Chemother 60:7415-7423.

4.	Sherrard LJ, Tai AS, Wee BA, Ramsay KA, Kidd TJ, Ben Zakour NL, Whiley DM, Beatson SA, Bell SC. 2017. Within-host whole genome analysis of an antibiotic resistant *Pseudomonas aeruginosa* strain sub-type in cystic fibrosis. PLoS ONE 12:e0172179.

5.	Ramanathan B, Jindal HM, Le CF, Gudimella R, Anwar A, Razali R, Poole-Johnson J, Manikam R, Sekaran SD. 2017. Next generation sequencing reveals the antibiotic resistant variants in the genome of *Pseudomonas aeruginosa*. PLoS ONE 12:e0182524.

6.	del Barrio-Tofiño E, López-Causapé C, Cabot G, Rivera A, Benito N, Segura C, Montero MM, Sorlí L, Tubau F, Gómez-Zorrilla S, Tormo N, Durá-Navarro R, Viedma E, Resino-Foz E, Fernández-Martínez M, González-Rico C, Alejo-Cancho I, Martínez JA, Labayru-Echverria C, Dueñas C, Ayestarán I, Zamorano L, Martinez-Martinez L, Horcajada JP, Oliver A. 2017. Genomics and susceptibility profiles of extensively drug-resistant *Pseudomonas aeruginosa* isolates from Spain. Antimicrob Agents Chemother 61:e01589-17.

7.	Centres for Disease Control and Prevention. 2022. CDC & FDA Antibiotic Resistance Isolate Bank. <https://wwwn.cdc.gov/ARIsolateBank/Panel/PanelDetail?ID=12>, accessed 18Jan22]. Accessed 1/18/2022.

8.	Buhl M, Kästle C, Geyer A, Autenrieth IB, Peter S, Willmann M. 2019. Molecular evolution of extensively drug-resistant (XDR) *Pseudomonas aeruginosa* strains from patients and hospital environment in a prolonged outbreak. Front Microbiol 10:1742.

9.	Wardell SJT, Rehman A, Martin LW, Winstanley C, Patrick WM, Lamont IL. 2019. A large-scale whole-genome comparison shows that experimental evolution in response to antibiotics predicts changes in naturally evolved clinical *Pseudomonas aeruginosa*. Antimicrob Agents Chemother 63:e01619-19.

10.	Khaledi A, Weimann A, Schniederjans M, Asgari E, Kuo T-H, Oliver A, Cabot G, Kola A, Gastmeier P, Hogardt M, Jonas D, Mofrad MR, Bremges A, McHardy AC, Häussler S. 2020. Predicting antimicrobial resistance in *Pseudomonas aeruginosa* with machine learning-enabled molecular diagnostics. EMBO Mol Med 12:e10264.

11.	Tsang KK, Maguire F, Zubyk HL, Chou S, Edalatmand A, Wright GD, Beiko RG, McArthur AG. 2021. Identifying novel β-lactamase substrate activity through *in silico* prediction of antimicrobial resistance. Microb Genom 7:mgen000500.

12.	Cortes-Lara S, Barrio-Tofino ED, Lopez-Causape C, Oliver A, Group G-SRPs. 2021. Predicting Pseudomonas aeruginosa susceptibility phenotypes from whole genome sequence resistome analysis. Clin Microbiol Infect 27:1631-1637.

13.	Sun Z, Yang F, Ji J, Cao W, Liu C, Ding B, Xu X. 2023. Dissecting the genotypic features of a fluoroquinolone-resistant Pseudomonas aeruginosa ST316 sublineage causing ear infections in Shanghai, China. Microb Genom 9.
