## Supplementary material for "Keeping up with the pathogens: Improved antimicrobial resistance detection and prediction from *Pseudomonas* aeruginosa genomes": Table S2

**Table S2.** *Pseudomonas aeruginosa* Validation Dataset strains generated in this study, and their corresponding antimicrobial susceptibility profiles.

| **Strain ID** | **Disease** | **Year of isolation** | **Antimicrobial sensitivity testing** | | | | | | | | | |
| --- | --- | --- | --- | --- | --- | --- | --- | --- | --- | --- | --- | --- |
|  |  |  | **AMK** | **TOB** | **PIP** | **TZP** | **CAZ** | **FEP** | **CIP** | **IPM** | **MEM** | **CST** |
| SCHI0002.S.8 | CF | 2017 | S | S | R | I | I | R | R | R | S | S |
| SCHI0002.S.9 | CF | 2017 | R | S | R | R | R | R | I | R | R | S |
| SCHI0002.S.12 | CF | 2018 | R | R | R | R | R | R | I | S | S | S |
| SCHI0003.S.1 | CF | 2017 | R | I | R | S | S | S | I | I | S | S |
| SCHI0003.S.3 | CF | 2017 | R | I | R | R | S | R | I | I | S | S |
| SCHI0003.S.8 | CF | 2017 | R | R | R | R | R | R | R | R | R | S |
| SCHI0004.S.5 | CF | 2017 | R | R | R | R | R | R | R | R | I | S |
| SCHI0005.S.8 | CF | 2017 | R | R | R | I | R | R | S | R | R | S |
| SCHI0005.S.9 | CF | 2017 | R | I | R | R | R | R | R | R | R | S |
| SCHI0005.S.10 | CF | 2017 | R | I | R | R | R | R | R | R | R | S |
| SCHI0005.S.11 | CF | 2017 | R | I | R | R | R | R | R | R | R | S |
| SCHI0006.S.10 | CF | 2017 | R | R | R | I | R | R | R | R | R | S |
| SCHI0008.S.2 | CF | 2018 | R | R | S | S | S | S | I | R | S | S |
| SCHI0008.S.3 | CF | 2018 | R | S | S | S | S | S | R | I | I | S |
| SCHI0008.S.4 | CF | 2018 | R | R | S | S | S | S | I | I | S | S |
| SCHI0010.S.1 | CF | 2018 | R | R | R | R | R | R | R | R | R | S |
| SCHI0013.S.2 | CF | 2018 | S | R | S | S | S | I | I | R | S | S |
| SCHI0013.S.12 | CF | 2018 | R | S | R | S | S | R | I | R | I | S |
| SCHI0016.S.57 | BE | 2017 | S | S | R | R | R | R | I | R | R | S |
| SCHI0016.S.124 | UTI | 2018 | R | S | R | I | I | I | R | R | R | S |
| SCHI0020.S.4 | CF | 2019 | S | S | R | S | S | S | S | S | S | S |
| SCHI0020.S.5 | CF | 2019 | R | S | R | S | S | I | S | S | S | S |
| SCHI0020.S.6 | CF | 2019 | R | S | S | S | S | R | S | S | S | S |
| SCHI0021.S.5 | CF | 2019 | R | R | R | S | S | R | I | S | S | S |
| SCHI0021.S.6 | CF | 2019 | R | R | R | S | S | R | R | S | S | S |
| SCHI0021.S.7 | CF | 2019 | R | R | S | S | S | R | R | S | S | S |
| SCHI0021.S.8 | CF | 2019 | R | R | S | S | S | R | R | S | S | S |
| SCHI0021.S.10 | CF | 2019 | R | I | S | S | S | R | I | S | S | S |
| SCHI0021.S.11 | CF | 2019 | R | R | S | S | S | R | R | S | S | S |
| SCHI0021.S.12 | CF | 2019 | R | R | S | S | S | R | R | S | S | S |
| SCHI0021.S.14 | CF | 2019 | R | R | S | S | S | R | R | S | S | S |
| SCHI0025.S.9 | CF | 2019 | R | R | R | R | I | R | I | R | R | S |
| SCHI0025.S.15 | CF | 2019 | R | R | R | R | R | R | R | R | R | S |
| SCHI0027.S.1 | CF | 2019 | R | R | R | R | R | R | R | R | R | S |
| SCHI0027.S.2 | CF | 2019 | R | R | R | R | R | R | R | R | R | S |
| SCHI0027.S.3 | CF | 2019 | R | R | R | R | R | R | R | R | R | S |
| SCHI0027.S.4 | CF | 2019 | R | I | S | S | S | S | S | S | S | S |
| SCHI0027.S.5 | CF | 2019 | R | I | R | R | R | R | R | R | R | S |
| SCHI0029.S.1 | CF | 2019 | R | R | R | R | R | R | R | R | R | S |
| SCHI0030.S.1 | CF | 2019 | S | S | S | S | S | I | S | S | S | S |
| SCHI0030.S.2 | CF | 2019 | R | R | R | R | R | R | R | R | R | S |
| SCHI0030.S.3 | CF | 2019 | R | I | R | R | R | R | R | R | R | S |
| SCHI0030.S.4 | CF | 2019 | R | R | R | R | R | R | R | R | R | S |
| SCHI0030.S.5 | CF | 2019 | S | S | S | S | S | S | S | S | S | S |
| SCHI0032.S.32 (AUS205) | Ulcer | 2008 | S | S | R | I | S | S | R | S | S | S |
| SCHI0032.S.33 (AUS134) | Ear Infection | 2008 | S | S | S | S | S | S | R | S | S | S |
| SCHI0033.S.1 | BSI | 2008 | S | S | R | R | R | R | S | S | S | S |
| SCHI0033.S.2 | BSI | 2009 | I | S | R | R | R | R | R | R | R | S |
| SCHI0033.S.3 | BSI | 2010 | S | S | R | S | S | S | S | S | S | S |
| SCHI0033.S.4 | BSI | 2010 | S | R | R | R | S | R | R | I | R | S |
| SCHI0033.S.5 | BSI | 2010 | S | S | R | R | R | R | S | S | S | S |
| SCHI0033.S.7 | BSI | 2010 | S | S | S | S | S | S | S | S | S | S |
| SCHI0033.S.8 | BSI | 2010 | S | S | R | R | R | R | S | S | S | S |
| SCHI0033.S.9 | BSI | 2010 | S | S | R | R | R | R | S | S | S | S |
| SCHI0033.S.10 | BSI | 2010 | S | S | R | R | R | I | S | S | S | S |
| SCHI0033.S.11 | BSI | 2012 | S | S | R | I | S | S | S | S | S | S |
| SCHI0033.S.12 | BSI | 2012 | R | S | R | I | R | R | S | S | S | S |
| SCHI0033.S.13A | BSI | 2012 | S | S | R | S | S | S | S | S | S | S |
| SCHI0033.S.13B | BSI | 2012 | S | S | S | S | S | S | S | S | S | S |
| SCHI0033.S.14 | BSI | 2008 | S | S | R | I | R | R | R | I | S | S |
| SCHI0033.S.15 | BSI | 2009 | R | R | R | I | R | R | R | R | R | S |
| SCHI0033.S.16 | BSI | 2009 | S | S | R | R | R | R | S | S | S | S |
| SCHI0033.S.17 | BSI | 2009 | S | S | S | S | S | S | S | S | S | S |
| SCHI0033.S.18 | BSI | 2010 | S | S | S | S | S | S | S | S | S | S |
| SCHI0033.S.19 | BSI | 2010 | R | R | R | S | R | R | R | S | S | S |
| SCHI0033.S.20 | BSI | 2010 | S | S | R | I | I | I | S | S | S | S |
| SCHI0033.S.23 | BSI | 2009 | S | S | R | S | S | S | S | S | I | S |
| SCHI0033.S.24 | BSI | 2009 | S | S | R | I | I | I | S | S | S | S |
| SCHI0033.S.25 | BSI | 2010 | S | S | S | S | S | S | S | R | R | S |
| SCHI0033.S.26 | BSI | 2010 | S | S | S | S | S | R | R | R | I | S |
| SCHI0033.S.27 | BSI | 2010 | S | S | R | I | I | R | S | S | S | S |
| SCHI0033.S.28 | BSI | 2010 | S | S | R | S | S | S | S | R | R | S |
| SCHI0033.S.29 | BSI | 2010 | S | S | S | S | S | S | I | S | S | S |
| SCHI0033.S.30 | BSI | 2010 | S | S | S | S | S | S | S | R | I | S |
| SCHI0033.S.32 | BSI | 2010 | I | S | S | S | S | S | S | R | I | S |
| SCHI0033.S.33 | BSI | 2010 | S | S | R | I | S | I | S | R | R | S |
| SCHI0033.S.35 | BSI | 2010 | S | S | S | S | S | S | S | S | S | S |
| SCHI0033.S.36 | BSI | 2012 | S | S | R | I | S | I | S | R | R | S |
| SCHI0033.S.37 | BSI | 2012 | S | S | S | S | S | S | S | I | S | S |
| SCHI0033.S.38 | BSI | 2012 | S | R | R | S | S | S | S | R | S | S |
| SCHI0033.S.39 | BSI | 2012 | S | S | S | S | S | S | S | I | S | S |
| SCHI0038.S.1 | COPD | 2020 | S | ND | R | S | S | S | S | S | S | S |
| SCHI0038.S.3 | COPD | 2020 | S | S | S | S | S | S | S | S | S | S |
| SCHI0039.S.1 | COPD | 2020 | S | S | S | S | S | S | S | S | S | S |
| SCHI0050.S.1 | COPD | 2020 | S | S | S | S | S | S | S | S | S | S |
| SCHI0050.S.3 | COPD | 2020 | S | ND | S | S | S | S | S | S | S | S |
| SCHI0058.S.1 | COPD | 2020 | S | S | S | S | S | S | S | S | S | S |
| SCHI0058.S.2 | COPD | 2021 | S | ND | S | S | S | S | S | S | S | S |
| SCHI0059.S.1 | COPD | 2020 | S | S | S | S | S | S | S | S | S | S |
| SCHI0064.S.1 | COPD | 2020 | S | S | S | S | S | S | S | S | S | S |
| SCHI0065.S.1 | COPD | 2020 | S | S | S | S | S | S | S | S | S | S |
| SCHI0068.S.3 | COPD | 2020 | S | ND | S | S | S | S | S | S | S | S |
| SCHI0070.S.1 | COPD | 2020 | S | S | S | S | S | S | S | S | S | S |
| SCHI0071.S.1 | COPD | 2020 | S | ND | S | S | S | S | S | S | S | S |
| SCHI0084.S.1 | COPD | 2021 | S | ND | R | S | S | S | S | S | S | S |
| SCHI0098.S.1 | COPD | 2021 | S | ND | S | S | S | S | R | S | S | S |
| SCHI0103.S.1 | COPD | 2021 | S | ND | S | S | S | S | S | S | S | S |
| SCHI0107.S.1 | COPD | 2022 | S | ND | S | S | S | S | S | S | S | S |
| SCHI0109.S.1 | COPD | 2021 | S | ND | R | S | S | S | S | S | S | S |
| SCHI0109.S.2 | COPD | 2022 | S | ND | R | R | R | I | R | S | S | S |
| SCHI0112.S.1 | COPD | 2021 | S | ND | S | S | S | S | S | S | S | S |
| SCHI0112.S.2 | COPD | 2022 | S | ND | S | S | S | S | S | S | S | S |

Abbreviations: AMK, amikacin; BE, bronchiectasis; BSI, blood stream infection; CAZ, ceftazidime; CF, cystic fibrosis; CIP, ciprofloxacin; COPD, chronic obstructive pulmonary disease; CST, colistin; FEP, cefepime; IPM, imipenem; MEM, meropenem; PIP, piperacillin; TOB, tobramycin; TZP, piperacillin/tazobactam; UTI, urinary tract infection.

Grey shading denotes intermediate (I) or resistant (R) phenotype for a given antibiotic. Non-grey-shaded cells denote a sensitive phenotype for a given antibiotic. ETESTs were used to determine minimum inhibitory concentrations for ciprofloxacin and meropenem. All other antibiotic resistance profiling was performed using disc diffusions.
