## Supplementary material for "Keeping up with the pathogens: Improved antimicrobial resistance detection and prediction from *Pseudomonas* aeruginosa genomes": Table S3

**Table S3.** List of naturally occurring chromosomal determinants not associated with conferring antimicrobial resistance in *Pseudomonas aeruginosa*, and thus ignored by ARDaP.

| **Locus** | **Mutation** | **Reference** |
| --- | --- | --- |
| *PA0004* (*gyrB*) | Ser466Phe | [1] |
| *PA0090* (*clpV1*) | LOF | [2] |
| *PA0227* | LOF | [2] |
| *PA0262* | LOF | This study |
| *PA0295* | Ala155Thr | This study |
| *PA0373* (*ftsY*) | Glu127_Pro130dup | This study |
| *PA0424* (*mexR*) | LOF | [3] |
| *PA0424* (*mexR*) | Ile24fs | This study |
| *PA0424* (*mexR*) | Gly71Glu | [3] |
| *PA0424* (*mexR*) | Ala103Thr | This study |
| *PA0424* (*mexR*) | Ala108fs | This study |
| *PA0424* (*mexR*) | Ile111fs | This study |
| *PA0424* (*mexR*) | Val126Glu | [3, 4] |
| *PA0424* (*mexR*) | Glu153Gln | [3] |
| *PA0424* (*mexR*) | Ser209Arg | [3] |
| *PA0425* (*mexA*) | Gly108Thr | [5] |
| *PA0425* (*mexA*) | Gln183* | This study |
| *PA0425* (*mexA*) | Tyr197* | [6] |
| *PA0425* (*mexA*) | Cys360Gly | [6] |
| *PA0426* (*mexB*) | Val45Leu | [7] |
| *PA0426* (*mexB*) | Gln773* | This study |
| *PA0426* (*mexB*) | Met901fs | [6] |
| *PA0427* (*oprM*) | LOF | [8] |
| *PA0575* | LOF | This study |
| *PA0650* (*trpD*) | LOF | This study |
| *PA0799* | Ala275Thr | This study |
| *PA0946* | Arg287Trp | This study |
| *PA0958* (*oprD*) | Asp43Asn | This study |
| *PA0958* (*oprD*) | Ser57fs | This study |
| *PA0958* (*oprD*) | Ser57Glu | [9, 10] |
| *PA0958* (*oprD*) | Ser59Arg | [9, 10] |
| *PA0958* (*oprD*) | Ser59fs | This study |
| *PA0958* (*oprD*) | Thr103Ser | [10, 11] |
| *PA0958* (*oprD*) | Lys115Thr | [11] |
| *PA0958* (*oprD*) | Val127Leu | [9] |
| *PA0958* (*oprD*) | Val153Leu | This study |
| *PA0958* (*oprD*) | Phe170Leu | [10, 11] |
| *PA0958* (*oprD*) | Glu185Gln | [9-11] |
| *PA0958* (*oprD*) | Pro186fs | This study |
| *PA0958* (*oprD*) | Pro186Gly | [10, 11] |
| *PA0958* (*oprD*) | Thr187fs | This study |
| *PA0958* (*oprD*) | Val189del | This study |
| *PA0958* (*oprD*) | Val189Thr | [10, 11] |
| *PA0958* (*oprD*) | Glu202Gln | [10] |
| *PA0958* (*oprD*) | Ile210Ala | [10] |
| *PA0958* (*oprD*) | Glu230Lys | [9, 10] |
| *PA0958* (*oprD*) | Ser240Thr | [9, 10] |
| *PA0958* (*oprD*) | Asn262Thr | [9, 10] |
| *PA0958* (*oprD*) | Gly265Ser | [10] |
| *PA0958* (*oprD*) | Ala267Ser | [9] |
| *PA0958* (*oprD*) | Thr276Ala | [12] |
| *PA0958* (*oprD*) | Ala281fs | This study |
| *PA0958* (*oprD*) | Ala281Gly | [10] |
| *PA0958* (*oprD*) | Ala282fs | This study |
| *PA0958 (oprD)* | Ala293Pro | [11] |
| *PA0958* (*oprD*) | Lys296Gln | [10] |
| *PA0958* (*oprD*) | Gln301Glu | [10] |
| *PA0958* (*oprD*) | Arg310Gly | [9] |
| *PA0958* (*oprD*) | Ala315Gly | [9, 11] |
| *PA0958* (*oprD*) | Leu347Ile | This study |
| *PA0958* (*oprD*) | Val359Leu | [9] |
| *PA0958* (*oprD*) | Met372fs | [11, 13] |
| *PA0958* (*oprD*) | Met372Val | [9] |
| *PA0958* (*oprD*) | Asp374dup | This study |
| *PA0958* (*oprD*) | Asp374fs | This study |
| *PA0958* (*oprD*) | Asn375Ser | [9] |
| *PA0958* (*oprD*) | Asn376fs | This study |
| *PA0958* (*oprD*) | Asn376Ser | [9] |
| *PA0958* (*oprD*) | Val377fs | This study |
| *PA0958* (*oprD*) | Gly378fs | [9] |
| *PA0958* (*oprD*) | Lys380_Asn381del | This study |
| *PA0958* (*oprD*) | Lys380fs | This study |
| *PA0958* (*oprD*) | Lys380Tyr | [9] |
| *PA0958* (*oprD*) | Asn381fs | This study |
| *PA0958* (*oprD*) | Asn381His | This study |
| *PA0958* (*oprD*) | Tyr382_Gly383insAla | This study |
| *PA0958* (*oprD*) | Tyr382Gly | [9] |
| *PA0958* (*oprD*) | Gly383_Tyr384insLeu | This study |
| *PA0958* (*oprD*) | Asn407Ala | This study |
| *PA0958* (*oprD*) | His418Ala | [9] |
| *PA0958* (*oprD*) | Arg419Pro | [9] |
| *PA0958* (*oprD*) | Ala420Cys | [9] |
| *PA0958* (*oprD*) | Asn421Gln | [9] |
| *PA0958* (*oprD*) | Ala422_Asp423del | [9] |
| *PA0958* (*oprD*) | Ala422Arg | [9] |
| *PA0958* (*oprD*) | Asp423Arg | [9] |
| *PA0958* (*oprD*) | Gln424Glu | [9] |
| *PA0958* (*oprD*) | Gln424Pro | [9] |
| *PA0958* (*oprD*) | Gly425Ala | [11] |
| *PA0958* (*oprD*) | Glu426Arg | [9] |
| *PA0958* (*oprD*) | Gly427Arg | [9] |
| *PA0958* (*oprD*) | Asp428Arg | [9] |
| *PA0958* (*oprD*) | Gln429Pro | [9] |
| *PA0958* (*oprD*) | Asn430Glu | [9] |
| *PA0958* (*oprD*) | Glu431Arg | [9] |
| *PA0958* (*oprD*) | Tyr438fs | This study |
| *PA1017* (*paua*) | LOF | This study |
| *PA1045* | Lys322Arg | This study |
| *PA1101* (*fliF*) | LOF | [2] |
| *PA1167* | LOF | [2] |
| *PA1171* | LOF | [2] |
| *PA1180 (phoQ)* | LOF | [14] |
| *PA1180* (*phoQ*) | Val260Gly | [15] |
| *PA1195* | LOF | [2] |
| *PA1259* | LOF | [2] |
| *PA1316* | LOF | [2] |
| *PA1375* (*pdxB*) | Glu367Gly | This study |
| *PA1430* (*lasR*) | Gly191Ser | This study |
| *PA1487* | Arg185Ser | This study |
| *PA1549* (*copA2*) | LOF | [2] |
| *PA1549* (*copA2*) | Ala441fs | [6] |
| *PA1549* (*copA2*) | Ala442fs | [6] |
| *PA1611* | LOF | [2] |
| *PA1798* (*parS*) | LOF | This study |
| *PA1798* (*parS*) | Leu137Pro | [16] |
| *PA1798* (*parS*) | Ala138Thr | [16] |
| *PA1798* (*parS*) | His398Arg | [17] |
| *PA1799* (*parR*) | Met59Ile | This study |
| *PA1997* | Ile54Val | This study |
| *PA2020* | LOF | [16] |
| *PA2020* | Asn54_Lys55dup | This study |
| *PA2023* (*galU*) | LOF | [2] |
| *PA2064* (*pcoB*) | Gln59Lys | This study |
| *PA2119* | LOF | This study |
| *PA2128* (*cupA1*) | LOF | This study |
| *PA2152* | LOF | This study |
| *PA2198* | LOF | [2] |
| *PA2207* | LOF | [2] |
| *PA2252* | LOF | This study |
| *PA2326* | LOF | [2] |
| *PA2491 (mexS)* | LOF | [18] |
| *PA2491 (mexS)* | Arg48Cys | [19] |
| *PA2491* (*mexS*) | Ser124Arg | [18] |
| *PA2491 (mexS)* | Asp249Asn | [20] |
| *PA2492* (*mexT*) | LOF | This study |
| *PA2492* (*mexT*) | Ala21fs | This study |
| *PA2492* (*mexT*) | Leu26Val | [21] |
| *PA2492* (*mexT*) | Arg28Ser | [22] |
| *PA2492* (*mexT*) | Ala39Val | [22] |
| *PA2492* (*mexT*) | Pro75fs | This study |
| *PA2492* (*mexT*) | Ala78fs | This study |
| *PA2492* (*mexT*) | Gln80fs | This study |
| *PA2492* (*mexT*) | Arg111fs | This study |
| *PA2492* (*mexT*) | Tyr164Asp | This study |
| *PA2492* (*mexT*) | Leu183fs | This study |
| *PA2492* (*mexT*) | Glu192fs | This study |
| *PA2492* (*mexT*) | Gly284Asp | This study |
| *PA2492* (*mexT*) | Ala293fs | This study |
| *PA2519* (*xylS*) | Thr242Pro | This study |
| *PA2571* | LOF | [2] |
| *PA2567* | Ile508Val | This study |
| *PA2638 (nuoB)* | LOF | This study |
| *PA2727* | Arg383Gln | This study |
| *PA2882* | Gln156His | This study |
| *PA2951* (*etfA*) | LOF | [2] |
| *PA2953* | LOF | [2] |
| *PA3015* | LOF | [2] |
| *PA3127* | LOF | [2] |
| *PA3168 (gyrA)* | Asp87Gly | [23, 24] |
| *PA3168* (*gyrA*) | Ala136Val | [24] |
| *PA3168* (*gyrA*) | Ala458Val | This study |
| *PA3178* | Ala115Val | This study |
| *PA3324* | LOF | [2] |
| *PA3351* (*flgM*) | LOF | [2] |
| *PA3418* (*ldh*) | LOF | This study |
| *PA3491* (*rnfC*) | Thr68Ser | This study |
| *PA3491* (*rnfC*) | His87Pro | [23] |
| *PA3491* (*rnfC*) | Cys198Arg | This study |
| *PA3491* (*rnfC*) | Ala639Val | [23] |
| *PA3491* (*rnfC*) | Gly652Arg | This study |
| *PA3491* (*rnfC*) | Ile726Thr | This study |
| *PA3491* (*rnfC*) | Ala751Pro | This study |
| *PA3491* (*rnfC*) | Ala759Thr | This study |
| *PA3574* | LOF | [2] |
| *PA3576* | LOF | [2] |
| *PA3574* (*nalD*) | Leu99fs | This study |
| *PA3574* (*nalD*) | Pro159Gln | This study |
| *PA3574* (*nalD*) | Asp187His | [25] |
| *PA3574 (nalD)* | Thr188Ala | [25, 26] |
| *PA3620 (mutS)* | LOF | [27] |
| *PA3622 (rpoS)* | LOF | This study |
| *PA3721* (*nalC*) | LOF | [6] |
| *PA3721* (*nalC*) | Thr50Pro | This study |
| *PA3721* (*nalC*) | Gly71Glu | [3, 21, 25, 28, 29] |
| *PA3721* (*nalC*) | Asp147Asn | [21] |
| *PA3721* (*nalC*) | Ala186Thr | [29] |
| *PA4020* (*mpl*) | Met38fs | [30] |
| *PA4020* (*mpl*) | Met297Val | This study |
| *PA4020* (*mpl*) | Arg322Gln | [31] |
| *PA4020 (mpl)* | LOF | [32] |
| *PA4020* (*mpl*) | Ter452Argext*? | [32] |
| *PA4025* | LOF | [2] |
| *PA4105* | LOF | This study |
| *PA4109* (*ampR*) | LOF | This study |
| *PA4109* (*ampR*) | Ala51Thr | [33] |
| *PA4109* (*ampR*) | Glu114Ala | [33] |
| *PA4109* (*ampR*) | Arg244Trp | [21] |
| *PA4109* (*ampR*) | Gly273Glu | [21] |
| *PA4109* (*ampR*) | Gly283Glu | [21, 33] |
| *PA4109* (*ampR*) | Met288Arg | [21, 33] |
| *PA4109* (*ampR*) | Glu292fs | This study |
| *PA4109* (*ampR*) | Ala293fs | This study |
| *PA4109* (*ampR*) | Arg294fs | This study |
| *PA4109* (*ampR*) | Arg296fs | This study |
| *PA4110* (*ampC*) | Thr21Ala | [34] |
| *PA4110* (*ampC*) | Gly27Asp | [34] |
| *PA4110* (*ampC*) | Gly27Val | [35] |
| *PA4110* (*ampC*) | Ala55Thr | [35] |
| *PA4110* (*ampC*) | Arg79Gln | [34, 35] |
| *PA4110* (*ampC*) | Ala97Val | [25, 34] |
| *PA4110* (*ampC*) | Thr105Ala | [34, 35] |
| *PA4110* (*ampC*) | Lys108Glu | [35] |
| *PA4110* (*ampC*) | Gln155Arg | [35] |
| *PA4110* (*ampC*) | Leu176Arg | [35] |
| *PA4110* (*ampC*) | Gly186Ser | [34] |
| *PA4110* (*ampC*) | Met201Leu | [35] |
| *PA4110* (*ampC*) | Val205Leu | [35] |
| *PA4110* (*ampC*) | Val356Ile | [35] |
| *PA4110* (*ampC*) | Gly391Ala | [35] |
| *PA4120* | Ser158fs | This study |
| *PA4120* | Gly159fs | This study |
| *PA4218 (ampP)* | Met87Ile | [31] |
| *PA4218 (ampP)* | Arg171Cys | [31] |
| *PA4218 (ampP)* | Thr172Ala | [31] |
| *PA4266* (*fusA1*) | Ala21Val | [6] |
| *PA4266* (*fusA1*) | Val93Ala | [36] |
| *PA4266* (*fusA1*) | Ile186Val | [36] |
| *PA4293 (pprA)* | LOF | [37] |
| *PA4310 (pctB)* | LOF | This study |
| *PA4315 (mvtA)* | LOF | [20] |
| *PA4334* | LOF | This study |
| *PA4375* | Gly930Glu | This study |
| *PA4418* (*ftsI*) | Pro215Leu | [38, 39] |
| *PA4418* (*ftsI*) | Gly216Ser | [38] |
| *PA4490* | LOF | [2] |
| *PA4521 (ampE)* | Ser69Pro | [31] |
| *PA4522* (*ampD*) | Asp28Gly | [40] |
| *PA4522* (*ampD*) | Gly46Ser | This study |
| *PA4522* (*ampD*) | His98Arg | This study |
| *PA4522* (*ampD*) | Ala136Val | [41] |
| *PA4522* (*ampD*) | Glu148Ala | [31] |
| *PA4522* (*ampD*) | Ser175Leu | [41] |
| *PA4522* (*ampD*) | Asp183Tyr | [31] |
| *PA4539* | LOF | This study |
| *PA4556* (*pilE*) | Leu121fs | This study |
| *PA4598* (*mexD*) | LOF | This study |
| *PA4599* (*mexC*) | -67C>T | This study |
| *PA4600* (*nfxB*) | Arg21His | [42] |
| *PA4600* (*nfxB*) | Asp56Gly | [43] |
| *PA4600* (*nfxB*) | Arg82Leu | [44] |
| *PA4600 (nfxB)* | LOF | [24] |
| *PA4622* | Ile380Val | This study |
| *PA4777* (*pmrB*) | Val15Ile | [45] |
| *PA4777* (*pmrB*) | Ala67Thr | [45] |
| *PA4777* (*pmrB*) | Asp70Asn | [45] |
| *PA4777* (*pmrB*) | Met292Ile | [46, 47] |
| *PA4777* (*pmrB*) | His340Arg | [45] |
| *PA4777* (*pmrB*) | Thr343Ala | [45] |
| *PA4777* (*pmrB*) | Tyr345His | [48] |
| *PA4778* (*cueR*) | Gly57Ala | This study |
| *PA4781* | LOF | [2] |
| *PA4898 (opdK)* | LOF | This study |
| *PA4946 (mutL)* | LOF | [27] |
| *PA4964 (parC)* | Gln405Arg | This study |
| *PA4964* (*parC*) | Val646Leu | This study |
| *PA4967* (*parE*) | Met437Ile | This study |
| *PA4967* (*parE*) | Ala473Val | [49-51] |
| *PA4967* (*parE*) | Asp533Glu | This study |
| *PA5001* | LOF | [2, 52] |
| *PA5003* | LOF | [2, 52] |
| *PA5003* | Arg268* | This study |
| *PA5038* (*aroB*) | Val85Ala | [7] |
| *PA5038* (*aroB*) | Ala200Glu | [7] |
| *PA5038* (*aroB*) | Thr211fs | This study |
| *PA5087* | Leu169Val | This study |
| *PA5114* | LOF | [2] |
| *PA5192 (pckA*) | LOF | [53] |
| *PA5192* (*pckA*) | Ala461Thr | This study |
| *PA5238* | Val85fs | This study |
| *PA5253* (*algP*) | LOF | This study |
| *PA5443* (*uvrD*) | LOF | [27] |
| *PA5471* (*armZ*) | Upregulation | [16] |
| *PA5485* (*ampDh2*) | -12_-11delTT | This study |
| *PA5528* | LOF | [54] |

Abbreviations: fs, frameshift; LOF, loss of function; *, stop codon. NB. Natural variation, including LOF mutations, does not exclude a gene as having a role in conferring antimicrobial resistance.

**References**

1.	Bruchmann S, Dötsch A, Nouri B, Chaberny IF, Häussler S: **Quantitative contributions of target alteration and decreased drug accumulation to *Pseudomonas aeruginosa* fluoroquinolone resistance**. *Antimicrob Agents Chemother* 2013, **57**(3):1361-1368.

2.	Dötsch A, Becker T, Pommerenke C, Magnowska Z, Jänsch L, Häussler S: **Genomewide identification of genetic determinants of antimicrobial drug resistance in *Pseudomonas aeruginosa***. *Antimicrob Agents Chemother* 2009, **53**(6):2522-2531.

3.	Llanes C, Hocquet D, Vogne C, Benali-Baitich D, Neuwirth C, Plésiat P: **Clinical strains of *Pseudomonas aeruginosa* overproducing MexAB-OprM and MexXY efflux pumps simultaneously**. *Antimicrob Agents Chemother* 2004, **48**(5):1797-1802.

4.	Nguyen KV, Nguyen TV, Nguyen HTT, Le DV: **Mutations in the *gyrA*, *parC*, and *mexR* genes provide functional insights into the fluoroquinolone-resistant *Pseudomonas aeruginosa* isolated in Vietnam**. *Infect Drug Resist* 2018, **11**:275-282.

5.	Chalhoub H, Sáenz Y, Nichols WW, Tulkens PM, Van Bambeke F: **Loss of activity of ceftazidime-avibactam due to MexAB-OprM efflux and overproduction of AmpC cephalosporinase in *Pseudomonas aeruginosa* isolated from patients suffering from cystic fibrosis**. *Int J Antimicrob Agents* 2018, **52**(5):697-701.

6.	Wardell SJT, Rehman A, Martin LW, Winstanley C, Patrick WM, Lamont IL: **A large-scale whole-genome comparison shows that experimental evolution in response to antibiotics predicts changes in naturally evolved clinical *Pseudomonas aeruginosa***. *Antimicrob Agents Chemother* 2019, **63**(12):e01619-01619.

7.	Cabot G, López-Causapé C, Ocampo-Sosa AA, Sommer LM, Domínguez MÁ, Zamorano L, Juan C, Tubau F, Rodríguez C, Moyà B *et al*: **Deciphering the resistome of the widespread *Pseudomonas aeruginosa* sequence type 175 international high-risk clone through whole-genome sequencing**. *Antimicrobial Agents and Chemotherapy* 2016, **60**(12):7415-7423.

8.	Wang H, Meng J, Jia M, Ma X, He G, Yu J, Wang R, Bai H, Hou Z, Luo X: **oprM as a new target for reversion of multidrug resistance in Pseudomonas aeruginosa by antisense phosphorothioate oligodeoxynucleotides**. *FEMS Immunol Med Microbiol* 2010, **60**(3):275-282.

9.	González-Vázquez MC, Rocha-Gracia RDC, Carabarín-Lima A, Bello-López E, Huerta-Romano F, Martínez-Laguna Y, Lozano-Zarain P: **Location of OprD porin in *Pseudomonas aeruginosa* clinical isolates**. *APMIS* 2021, **129**(4):213-224.

10.	Shu JC, Kuo AJ, Su LH, Liu TP, Lee MH, Su IN, Wu TL: **Development of carbapenem resistance in *Pseudomonas aeruginosa* is associated with OprD polymorphisms, particularly the amino acid substitution at codon 170**. *J Antimicrob Chemother* 2017, **72**(9):2489-2495.

11.	Rodríguez-Martínez JM, Poirel L, Nordmann P: **Molecular epidemiology and mechanisms of carbapenem resistance in *Pseudomonas aeruginosa***. *Antimicrobial Agents and Chemotherapy* 2009, **53**(11):4783-4788.

12.	Ocampo-Sosa AA, Cabot G, Rodríguez C, Roman E, Tubau F, Macia MD, Moya B, Zamorano L, Suárez C, Peña C *et al*: **Alterations of OprD in carbapenem-intermediate and -susceptible strains of *Pseudomonas aeruginosa* isolated from patients with bacteremia in a Spanish multicenter study**. *Antimicrob Agents Chemother* 2012, **56**(4):1703-1713.

13.	Epp SF, Köhler T, Plésiat P, Michéa-Hamzehpour M, Frey J, Pechère JC: **C-terminal region of *Pseudomonas aeruginosa* outer membrane porin OprD modulates susceptibility to meropenem**. *Antimicrob Agents Chemother* 2001, **45**(6):1780-1787.

14.	Lu L, Akerbladh L, Ahmad S, Konda V, Cao S, Vocat A, Maes L, Cole ST, Hughes D, Larhed M *et al*: **Synthesis and In Vitro Biological Evaluation of Quinolinyl Pyrimidines Targeting Type II NADH-Dehydrogenase (NDH-2)**. *ACS Infect Dis* 2022, **8**(3):482-498.

15.	Barrow K, Kwon DH: **Alterations in two-component regulatory systems of *phoPQ* and *pmrAB* are associated with polymyxin B resistance in clinical isolates of *Pseudomonas aeruginosa***. *Antimicrob Agents Chemother* 2009, **53**(12):5150-5154.

16.	Guénard S, Muller C, Monlezun L, Benas P, Broutin I, Jeannot K, Plésiat P: **Multiple mutations lead to MexXY-OprM-dependent aminoglycoside resistance in clinical strains of *Pseudomonas aeruginosa***. *Antimicrob Agents Chemother* 2014, **58**(1):221-228.

17.	Lee JY, Chung ES, Na IY, Kim H, Shin D, Ko KS: **Development of colistin resistance in *pmrA*-, *phoP*-, *parR*- and *cprR*-inactivated mutants of *Pseudomonas aeruginosa***. *J Antimicrob Chemother* 2014, **69**(11):2966-2971.

18.	Sobel ML, Neshat S, Poole K: **Mutations in *PA2491* (*mexS*) promote MexT-dependent *mexEF-oprN* expression and multidrug resistance in a clinical strain of *Pseudomonas aeruginosa***. *J Bacteriol* 2005, **187**(4):1246-1253.

19.	Petitjean M, Martak D, Silvant A, Bertrand X, Valot B, Hocquet D: **Genomic characterization of a local epidemic Pseudomonas aeruginosa reveals specific features of the widespread clone ST395**. *Microbial Genomics* 2017, **3**(10).

20.	Llanes C, Köhler T, Patry I, Dehecq B, van Delden C, Plésiat P: **Role of the MexEF-OprN efflux system in low-level resistance of *Pseudomonas aeruginosa* to ciprofloxacin**. *Antimicrob Agents Chemother* 2011, **55**(12):5676-5684.

21.	Quale J, Bratu S, Gupta J, Landman D: **Interplay of efflux system, *ampC*, and *oprD* expression in carbapenem resistance of *Pseudomonas aeruginosa* clinical isolates**. *Antimicrob Agents Chemother* 2006, **50**(5):1633-1641.

22.	LoVullo ED, Schweizer HP: ***Pseudomonas aeruginosa mexT* is an indicator of PAO1 strain integrity**. *J Med Microbiol* 2020, **69**(1):139-145.

23.	Rehman A, Jeukens J, Levesque RC, Lamont IL: **Gene-gene interactions dictate ciprofloxacin resistance in *Pseudomonas aeruginosa* and facilitate prediction of resistance phenotype from genome sequence data**. *Antimicrob Agents Chemother* 2021, **65**(7):e0269620.

24.	Higgins PG, Fluit AC, Milatovic D, Verhoef J, Schmitz FJ: **Mutations in GyrA, ParC, MexR and NfxB in clinical isolates of *Pseudomonas aeruginosa***. *International journal of antimicrobial agents* 2003, **21**(5):409-413.

25.	Tomás M, Doumith M, Warner M, Turton JF, Beceiro A, Bou G, Livermore DM, Woodford N: **Efflux pumps, OprD porin, AmpC beta-lactamase, and multiresistance in *Pseudomonas aeruginosa* isolates from cystic fibrosis patients**. *Antimicrob Agents Chemother* 2010, **54**(5):2219-2224.

26.	Quale J, Bratu S, Gupta J, Landman D: **Interplay of efflux system, *ampC*, and *oprD* expression in carbapenem resistance of *Pseudomonas aeruginosa* clinical isolates**. *Antimicrob Agents Chemother* 2006, **50**(5):1633-1641.

27.	Oliver A, Baquero F, Blázquez J: **The mismatch repair system (*mutS*, *mutL* and *uvrD* genes) in *Pseudomonas aeruginosa*: molecular characterization of naturally occurring mutants**. *Molecular microbiology* 2002, **43**(6):1641-1650.

28.	Díaz-Ríos C, Hernández M, Abad D, Álvarez-Montes L, Varsaki A, Iturbe D, Calvo J, Ocampo-Sosa AA: **New sequence type ST3449 in multidrug-resistant *Pseudomonas aeruginosa* isolates from a cystic fibrosis patient**. *Antibiotics (Basel)* 2021, **10**(5).

29.	Braz VS, Furlan JP, Fernandes AF, Stehling EG: **Mutations in NalC induce MexAB-OprM overexpression resulting in high level of aztreonam resistance in environmental isolates of *Pseudomonas aeruginosa***. *FEMS Microbiol Lett* 2016, **363**(16).

30.	Wang K, Chen YQ, Salido MM, Kohli GS, Kong JL, Liang HJ, Yao ZT, Xie YT, Wu HY, Cai SQ *et al*: **The rapid *in vivo* evolution of *Pseudomonas aeruginosa* in ventilator-associated pneumonia patients leads to attenuated virulence**. *Open Biol* 2017, **7**(9).

31.	Li Z, Cai Z, Cai Z, Zhang Y, Fu T, Jin Y, Cheng Z, Jin S, Wu W, Yang L *et al*: **Molecular genetic analysis of an XDR *Pseudomonas aeruginosa* ST664 clone carrying multiple conjugal plasmids**. *J Antimicrob Chemother* 2020, **75**(6):1443-1452.

32.	Tsutsumi Y, Tomita H, Tanimoto K: **Identification of novel genes responsible for overexpression of *ampC* in *Pseudomonas aeruginosa* PAO1**. *Antimicrob Agents Chemother* 2013, **57**(12):5987-5993.

33.	Balasubramanian D, Kumari H, Mathee K: ***Pseudomonas aeruginosa* AmpR: an acute-chronic switch regulator**. *Pathog Dis* 2015, **73**(2):1-14.

34.	Tam VH, Schilling AN, LaRocco MT, Gentry LO, Lolans K, Quinn JP, Garey KW: **Prevalence of AmpC over-expression in bloodstream isolates of *Pseudomonas aeruginosa***. *Clin Microbiol Infect* 2007, **13**(4):413-418.

35.	Berrazeg M, Jeannot K, Ntsogo Enguéné VY, Broutin I, Loeffert S, Fournier D, Plésiat P: **Mutations in β-Lactamase AmpC increase resistance of *Pseudomonas aeruginosa* isolates to antipseudomonal cephalosporins**. *Antimicrobial Agents and Chemotherapy* 2015, **59**(10):6248-6255.

36.	Bolard A, Plésiat P, Jeannot K: **Mutations in gene *fusA1* as a novel mechanism of aminoglycoside resistance in clinical strains of *Pseudomonas aeruginosa***. *Antimicrob Agents Chemother* 2018, **62**(2).

37.	Sherrard LJ, Tai AS, Wee BA, Ramsay KA, Kidd TJ, Ben Zakour NL, Whiley DM, Beatson SA, Bell SC: **Within-host whole genome analysis of an antibiotic resistant *Pseudomonas aeruginosa* strain sub-type in cystic fibrosis**. *PLoS ONE* 2017, **12**(3):e0172179.

38.	López-Causapé C, Sommer LM, Cabot G, Rubio R, Ocampo-Sosa AA, Johansen HK, Figuerola J, Cantón R, Kidd TJ, Molin S *et al*: **Evolution of the *Pseudomonas aeruginosa* mutational resistome in an international cystic fibrosis clone**. *Sci Rep* 2017, **7**(1):5555.

39.	McLean K, Lee D, Holmes EA, Penewit K, Waalkes A, Ren M, Lee SA, Gasper J, Manoil C, Salipante SJ: **Genomic analysis identifies novel *Pseudomonas aeruginosa* resistance genes under selection during inhaled aztreonam therapy *in vivo***. *Antimicrob Agents Chemother* 2019, **63**(9).

40.	Tueffers L, Barbosa C, Bobis I, Schubert S, Höppner M, Rühlemann M, Franke A, Rosenstiel P, Friedrichs A, Krenz-Weinreich A *et al*: ***Pseudomonas aeruginosa* populations in the cystic fibrosis lung lose susceptibility to newly applied β-lactams within 3 days**. *J Antimicrob Chemother* 2019, **74**(10):2916-2925.

41.	Langaee TY, Gagnon L, Huletsky A: **Inactivation of the *ampD* gene in *Pseudomonas aeruginosa* leads to moderate-basal-level and hyperinducible AmpC β-lactamase expression**. *Antimicrobial Agents and Chemotherapy* 2000, **44**(3):583-589.

42.	Jeannot K, Elsen S, Köhler T, Attree I, van Delden C, Plésiat P: **Resistance and virulence of *Pseudomonas aeruginosa* clinical strains overproducing the MexCD-OprJ efflux pump**. *Antimicrob Agents Chemother* 2008, **52**(7):2455-2462.

43.	Campo Esquisabel AB, Rodríguez MC, Campo-Sosa AO, Rodríguez C, Martínez-Martínez L: **Mechanisms of resistance in clinical isolates of *Pseudomonas aeruginosa* less susceptible to cefepime than to ceftazidime**. *Clin Microbiol Infect* 2011, **17**(12):1817-1822.

44.	Jalal S, Ciofu O, Hoiby N, Gotoh N, Wretlind B: **Molecular mechanisms of fluoroquinolone resistance in *Pseudomonas aeruginosa* isolates from cystic fibrosis patients**. *Antimicrob Agents Chemother* 2000, **44**(3):710-712.

45.	Lee JY, Ko KS: **Mutations and expression of PmrAB and PhoPQ related with colistin resistance in *Pseudomonas aeruginosa* clinical isolates**. *Diagn Microbiol Infect Dis* 2014, **78**(3):271-276.

46.	Abraham N, Kwon DH: **A single amino acid substitution in PmrB is associated with polymyxin B resistance in clinical isolate of *Pseudomonas aeruginosa***. *FEMS Microbiol Lett* 2009, **298**(2):249-254.

47.	Moskowitz SM, Brannon MK, Dasgupta N, Pier M, Sgambati N, Miller AK, Selgrade SE, Miller SI, Denton M, Conway SP *et al*: **PmrB mutations promote polymyxin resistance of *Pseudomonas aeruginosa* isolated from colistin-treated cystic fibrosis patients**. *Antimicrob Agents Chemother* 2012, **56**(2):1019-1030.

48.	Owusu-Anim D, Kwon DH: **Differential role of two-component regulatory cystems (*phoPQ* and *pmrAB*) in polymyxin B susceptibility of *Pseudomonas aeruginosa***. *Adv Microbiol* 2012, **2**(1).

49.	Akasaka T, Tanaka M, Yamaguchi A, Sato K: **Type II topoisomerase mutations in fluoroquinolone-resistant clinical strains of *Pseudomonas aeruginosa* isolated in 1998 and 1999: role of target enzyme in mechanism of fluoroquinolone resistance**. *Antimicrob Agents Chemother* 2001, **45**(8):2263-2268.

50.	Lee JK, Lee YS, Park YK, Kim BS: **Alterations in the GyrA and GyrB subunits of topoisomerase II and the ParC and ParE subunits of topoisomerase IV in ciprofloxacin-resistant clinical isolates of *Pseudomonas aeruginosa***. *Int J Antimicrob Agents* 2005, **25**(4):290-295.

51.	Rehman A, Jeukens J, Levesque RC, Lamont IL: **Gene-gene interactions dictate ciprofloxacin resistance in *Pseudomonas aeruginosa* and facilitate prediction of resistance phenotype from genome sequence data**. *Antimicrobial Agents and Chemotherapy* 2021.

52.	Alvarez-Ortega C, Wiegand I, Olivares J, Hancock RE, Martinez JL: **Genetic determinants involved in the susceptibility of Pseudomonas aeruginosa to beta-lactam antibiotics**. *Antimicrob Agents Chemother* 2010, **54**(10):4159-4167.

53.	Sun E, Gill EE, Falsafi R, Yeung A, Liu S, Hancock REW: **Broad-spectrum adaptive antibiotic resistance associated with *Pseudomonas aeruginosa* mucin-dependent surfing motility**. *Antimicrob Agents Chemother* 2018, **62**(9).

54.	Khaledi A, Weimann A, Schniederjans M, Asgari E, Kuo T-H, Oliver A, Cabot G, Kola A, Gastmeier P, Hogardt M *et al*: **Predicting antimicrobial resistance in *Pseudomonas aeruginosa* with machine learning-enabled molecular diagnostics**. *EMBO Mol Med* 2020, **12**(3):e10264-e10264.
