## Supplementary material for "Keeping up with the pathogens: Improved antimicrobial resistance detection and prediction from *Pseudomonas* aeruginosa genomes": Table S4

**Table S4.** Antimicrobial resistance rates across the Validation Dataset (*n*=102).

| **Antibiotic Class** | **Antibiotic** | **Resistant (%)** | **Intermediate (%)** | **Sensitive (%)** |
| --- | --- | --- | --- | --- |
| Carbapenems | MEM | 27 (26) | 7 (7) | 68 (67) |
|  | IPM | 35 (34) | 8 (8) | 59 (58) |
| Polymyxins | CST | 0 (0) | 0 (0) | 102 (100) |
| Fluoroquinolones | CIP | 35 (34) | 13 (13) | 54 (53) |
| Cephalosporins | FEP | 46 (45) | 10 (10) | 46 (45) |
|  | CAZ | 32 (31) | 6 (6) | 64 (63) |
| Penicillins | TZP | 29 (28) | 14 (14) | 59 (58) |
|  | PIP | 58 (57) | 0 (0) | 44 (43) |
| Aminoglycosides | TOB | 28 (31) | 9 (10) | 52 (58) |
|  | AMK | 41 (40) | 2 (2) | 59 (55) |

MEM, meropenem; IPM, imipenem; CST, colistin; CIP, ciprofloxacin; FEP, cefepime; CAZ, ceftazidime, TZP, piperacillin/tazobactam; PIP, piperacillin; TOB, tobramycin; AMK, amikacin
