## Supplementary material for "Keeping up with the pathogens: Improved antimicrobial resistance detection and prediction from *Pseudomonas* aeruginosa genomes": Table S5

**Table S5.** Assessment of ARDaP performance across the Global Dataset when including isolates with an intermediate resistance phenotype.

| **Antibiotic Class** | **Antibiotic** | **True Positive** | **True Negative** | **False Positive** | **False Negative** | **Specificity** | **Sensitivity** | **bACC** |
| --- | --- | --- | --- | --- | --- | --- | --- | --- |
| Carbapenems | MEM | 563 | 991 | 133 | 188 | 0.88 | 0.75 | 0.82 |
|  | IPM | 144 | 405 | 14 | 84 | 0.96 | 0.63 | 0.79 |
| Polymyxins | CST | 10 | 933 | 0 | 38 | 1.00 | 0.21 | 0.60 |
| Fluoroquinolones | CIP | 627 | 430 | 47 | 80 | 0.85 | 0.88 | 0.86 |
| Cephalosporins | FEP | 127 | 617 | 33 | 96 | 0.90 | 0.54 | 0.72 |
|  | CAZ | 255 | 690 | 43 | 119 | 0.90 | 0.63 | 0.77 |
| Penicillins | TZP | 112 | 504 | 22 | 81 | 0.93 | 0.52 | 0.72 |
|  | PIP | 154 | 255 | 17 | 41 | 0.91 | 0.77 | 0.84 |
| Aminoglycosides | TOB | 271 | 675 | 20 | 23 | 0.97 | 0.90 | 0.93 |
|  | AMK | 143 | 1158 | 39 | 48 | 0.94 | 0.72 | 0.83 |

Abbreviations: MEM, meropenem; IPM, imipenem; CST, colistin; CIP, ciprofloxacin; FEP, cefepime; CAZ, ceftazidime, TZP, piperacillin/tazobactam; PIP, piperacillin; TOB, tobramycin; AMK, amikacin; bACC, balanced accuracy.
